# Validation of an instrumented shoe insole framework for analyzing spatiotemporal gait metrics in healthy and neurodegenerative populations

**DOI:** 10.1101/2025.05.06.25326646

**Authors:** Matthew P. Mavor, Alexandre Mir-Orefice, Victor C.H. Chan, Gauruv Bose, Heather J. Maclean, Tiago Mestre, David Grimes, Mark S. Freedman, Ryan B. Graham

## Abstract

Many neurological conditions negatively affect a person’s walking quality, which is a vital aspect of their quality of life. Gait quality, through the collection of spatiotemporal variables, can also help infer disease status; however, in-clinic access to these metrics is limited or cannot be assessed frequently enough to proactively monitor disease progression (i.e., improvement, maintenance, worsening). To address these limitations, we developed a framework that analyzes spatiotemporal gait metrics using healthy and neurodegenerative walking data collected from instrumented shoe insoles. The Insole Framework (IF) identifies ambulatory activities using an artificial neural network, identifies gait events using logic, fuses the inertial measurement unit (IMU) data, standardizes the analysis to every ten seconds, and calculates spatiotemporal metrics categorized into core, pace, percentage, and asymmetry metrics. Activity classification algorithms had excellent accuracy and F1-score (≥ 93%). The spatiotemporal metrics obtained from the IF were validated against a gold standard motion capture system using ICCs, limits of agreement, and statistical testing. All core and pace metrics had good to excellent reliability and acceptable bias compared to the motion capture system, regardless of neurological function. Of the 19 spatiotemporal metrics assessed, system-independent statistical tests showed that similar population-level interpretations (i.e., one disagreement) and post-hoc differences (i.e., three disagreements) with similar levels of explained variance (absolute η^2^ difference between systems across all tests was 0.046) would be found regardless of the system used. The IF was considered valid and can appropriately capture ambulatory activities and spatiotemporal gait metrics in healthy, multiple sclerosis, and Parkinson’s disease populations.

**Author Summary:** Gait assessments are used by clinicians to infer the severity and progression of neurological diseases. These assessments aim to quantify gross walking quality (i.e., patient perception, visual observations, speed, and distance) rather than the spatiotemporal metrics (e.g., double support time, stride length, cadence, etc.) that differentiate people from controls, conditions, and severity levels. Although spatiotemporal metrics can be powerful digital biomarkers to assess disease severity and monitor progression, traditional motion capture methods are limited due to high costs, the need for specialized expertise, time-consuming analysis/operations and infrequent patient collections. To overcome these limitations, we propose a framework that uses instrumented shoe insoles (inertial measurement unit + pressure) to identify activities and analyze gait. With our framework, gait assessments can be done several times a month in free-living conditions instead of infrequent clinical gait assessments, reducing healthcare barriers and promoting objective decision-making. This work describes our activity recognition, gait detection, and fusion methods and demonstrates our framework’s ability to produce results comparable to a gold-standard motion capture system in participants with multiple sclerosis, Parkinson’s disease, and healthy individuals. Our Insole Framework is deemed valid due to high reliability, similar between-group interpretations across systems, and the activity recognition algorithm’s performance.

## 1.0 Introduction

A reduction in gait quality is one of the most common symptoms experienced by people with multiple sclerosis (1) (PwMS) and people with Parkinson’s disease (2) (PwPD) and is perceived as having the greatest impact on quality of life (3). This phenomenon has been exploited by clinical walking tests to infer disease progression by evaluating gross walking ability, such as total distance travelled over a set time (e.g., 6-minute walk test (4,5)) or distance (e.g., 500-metre walk test (6)) and time to complete set tasks (e.g., timed up and go (7)) or distances (e.g., 10-metre walk test, timed 25-foot walk (8,9)). However, by calculating spatiotemporal gait metrics, far more information can be gathered about the human gait pattern and, thus, inference about disease progression even in those who are at the earliest stages of neurological dysfunction (10).

Although spatiotemporal gait metrics can be powerful biomarkers to identify and track disease progression, the traditional tools needed to calculate these variables are mainly limited to laboratory environments and controlled protocols, limiting their widespread use and ecological validity (11). Even when clinicians have access to specialized laboratory equipment, the practical frequency of collecting these spatiotemporal gait metrics reduces the longitudinal benefit since minimal detectable changes need to occur to constitute clinical meaningfulness (12), signalling that irreversible changes to gait mechanics may have occurred between visits.

Rather than a reactive approach to healthcare, using wearable devices to capture subtle progressive changes in spatiotemporal gait metrics outside the clinic/laboratory in free-living environments multiple times a week would enable a proactive approach. Clinicians and patients alike would be able to identify early trends in walking quality (i.e., improvement, maintenance, worsening) to prescribe/advocate for early interventions (e.g., assistive devices, pharmacological, exercise, etc.) and continuously evaluate the impact of these interventions. However, there are several challenges to collecting spatiotemporal gait metrics outside a laboratory environment: the environment is unknown; ambulatory activities are vast and unpredictable; gait detection must be reliable regardless of ambulatory ability, disability status, assistive device usage, and environmental conditions; analyses and sensor placement must be standardized; and adherence is necessary for any long-term solution to be viable.

Instrumented shoe insoles are one type of wearable device that addresses many challenges with collecting longitudinal free-living gait data. These devices are invisible to users and non-users, have standard placements between data collections, and are easy for users to understand and use. Instrumented shoe insoles most commonly contain pressure sensors and an inertial measurement unit (IMU; (12), allowing for the development of precise gait detection algorithms that can leverage IMU-(13) and pressure-based (14,15) algorithms to identify key gait events. Researchers have validated spatiotemporal measures from commercially available insoles (16,17) and custom solutions (18), and have used them to discriminate between healthy older adults and people with Parkinson’s Disease (15). Instrumented insoles have also been used to identify dysfunctional gait patterns, such as shuffle gait (19), and have been leveraged by machine learning algorithms to detect gait events (20), activities of daily living (21–24), and spatiotemporal metrics (21). This evidence makes instrumented shoe insoles a viable choice for collecting long-term gait data in free-living conditions (25).

The purpose of this work was to validate an Insole Framework (IF) that uses pressure and IMU data obtained from instrumented shoe insoles to detect ambulatory activities to segment walking trials, perform gait detection, and calculate spatiotemporal gait metrics. The results obtained from our proposed framework are compared to data from a laboratory-based motion capture (MoCap) system using data collected from three populations: healthy participants (HP), PwMS, and PwPD.

## 2.0 Methods

### 2.1 Participants

Twenty-two HP, 19 PwMS, and 10 PwPD were recruited for this investigation. An HP was any individual who had not experienced a musculoskeletal injury within the last 6 months at the time of testing and did not suffer from a neurological disorder. PwMS and PwPD were recruited from The Ottawa Hospital and had undergone a neurological exam within the last 12 months, where they were given an Extended Disability Status Score (EDSS) up to 6.5 (PwMS; range: 0-10, with higher scores indicating higher levels of disability (6)) or a Hoehn and Yahr Scale (HY) score less than 3.0 (PwPD; range: 0-5, with higher scores indicating more severe disease progression(26)). Participants were asked to fill out the 12-item Multiple Sclerosis Walking Scale (MSWS-12) as a measure of perceived walking function (range: 0-100%, with higher values indicating increased walking impairment (27)); although designed for MS, PwPD were also asked to fill out the MSWS-12. The average EDSS for the PwMS was 3.8 ±1.6 (range: 0-6), the average MSWS-12 score was 60.9% ±22.2 (range: 20-90), and nine PwMS expressed using one or more assistive devices in their daily lives. The average HY score for the PwPD was 1.9 ±0.32 (range: 1-2), the average MSWS-12 score was 40.7% ±21.1 (range: 22-83), and no PwPD expressed daily usage of an assistive device. Participant demographics, disability metrics, and assistive device usage are presented in Table 1. Both the University of Ottawa (H-11-21-7565) and The Ottawa Health Science Network (20220239-01H) Research Ethics Boards approved this research.

**Table 1.** Participant Demographics.

|  | Sex<br>M\|F | Age<br>(years) | Height<br>(cm) | Mass<br>(kg) | EDSS<br>Score | MSWS-12 Score | MS Type | H-Y<br>Score | Assistive<br>device |
| --- | --- | --- | --- | --- | --- | --- | --- | --- | --- |
| Healthy | 11\|11 | 28.1<br>(6.66) | 174.8<br>(9.47) | 75.7<br>(14.8) | – | – | – | – | NAD: 22 |
| PwMS | 6\|13 | 54.2<br>(16.5) | 158.0<br>(7.28) | 78.7<br>(26.4) | 3.8 (1.6) | 60.9<br>(22.4) | RR: 10; PP:<br>4; SP: 1 | – | AFO: 1;<br>WS: 3;<br>W: 4; C:<br>5; NAD:<br>10 |
| PwPD | 6\|4 | 63.9<br>(7.19) | 159.1<br>(9.96) | 75.0<br>(15.1) | – | 40.7<br>(21.1) | – | 1.9<br>(0.32) | NAD: 10 |

Note: Values are presented as mean (standard deviation) or total count when appropriate. M = Male; F = Female, PwMS = People with Multiple Sclerosis; PwPD = People with Parkinson's Disease; MS = Multiple Sclerosis; EDSS = Expanded Disability Status Score; MSWS-12 = 12-Item Multiple Sclerosis Walking Scale; MS Types: RR = relapse remitting, PP = primary progressive, SP = secondary progressive; Assistive devices: AFO = ankle foot orthosis, WS = two walking sticks, W = walker, C = cane, NAD = no assistive device. Note, some PwMS expressed using multiple assistive devices (e.g., cane for short walks/with a companion and a walker for longer walks/using shopping carts), each mentioning of an assistive device was counted (i.e., for the previous example, cane and walker would each be counted as one instance). Four PwMS expressed using multiple assistive devices.

### 2.2 Movement Protocol and Instrumentation

Participants arrived at the University of Ottawa’s Movement Biomechanics and Analytics Laboratory for a single day. Following informed consent, PwMS and PwPD were asked to fill out the MSWS-12, which evaluated their perceived walking ability in the 2 weeks prior to data collection.(27) All participants were then asked to remove their indoor footwear so the appropriately sized pair of instrumented shoe insoles (ReGo, Moticon, Germany; 50 Hz) could be placed inside their shoes. The sizes used in this investigation ranged from S2 to S7 (EU shoe size 34–45, US shoe size Women 4-13, Men 3.5-11.5). For each walking trial, both instrumented shoe insoles streamed raw data from 16 pressure sensors and a triaxial accelerometer and gyroscope to a mobile application (Celestra Health, Canada) installed on a laboratory-owned smartphone (iPhone 13, Apple, USA). Walking tasks were performed in the laboratory (i.e., overground and treadmill walks), in the indoor hallways outside the laboratory (500- and 125-metre walks), and outdoors on a paved multiuse pathway (500-metre walk). Participants were asked to begin and end all walking trials by standing for approximately three seconds.

In the laboratory, all participants performed at least six overground walks measuring six metres over two force plates (FP-4060, Bertec, USA; 1000 Hz) placed in parallel. If participants arrived with an assistive device(s), they were asked if the six walks could be split evenly to progress them from unassisted to fully assisted walking (e.g., a participant with a cane would perform three unassisted walks and three with a cane). All healthy participants, optionally PwMS (N = 2) and PwPD (N = 0), performed a seven-minute walk on a treadmill at their preferred walking speed, determined as per Dingwell et al. (28) (i.e., iteratively speeding and slowing the treadmill to find the average preferred speed). Whole-body kinematics were collected using an eight-video-camera system (Vue, Vicon, UK; 50 Hz) and analyzed using Theia3D (Theia Markerless Inc., Canada), which has been validated for capturing gait metrics against marker-based motion capture systems (29).

In the hallways outside the laboratory, participants were asked to walk up to 500 metres around two pylons placed 25 metres apart; participants were asked to make alternating left- and right-hand turns around the pylons (i.e., making a figure 8). Participants were instructed to walk at a comfortable speed without stopping; a research assistant walked alongside participants at higher disability levels, and chairs were set up along the hallway at 5-metre intervals, allowing participants to stop and rest if needed. Participants were encouraged to perform the 500-metre walk without assistance; however, they were not prevented from using assistive devices. All healthy participants, optionally PwMS (N = 10) and PwPD (N = 9), also performed a 125-metre walk inside, which included several 90-180° turns in either direction, a stair ascent, and a stair descent section. Only healthy participants performed a 500-metre walk outside, which contained four ∼90° turns and a slight uphill and downhill section on a paved multiuse pathway.

Apart from the main protocol, a subset of healthy participants (N = 15) was invited to perform a stair ascent/descent protocol, where they walked up and down four flights of stairs twice under two conditions: assisted (i.e., actively using the railing to aid balance and propulsion) and preferred (i.e., no instructions given). This additional stair protocol was used to increase the training data for the activity recognition algorithm discussed below.

For all tasks outside the laboratory, participants were filmed by a researcher on a laboratory-owned smartphone to facilitate ground truth labelling for the human activity recognition (HAR) algorithm. Videos focused from below the neckline to the feet.

### 2.3 Framework Development

The framework developed to analyze gait patterns using instrumented shoe insole data first identified the ambulatory activities performed during the walking trial, detected gait phases, fused the IMU data to obtain foot position, standardized the analysis into 10-second segments, and calculated gait metrics.

#### 2.3.1 Human Activity Recognition (HAR)

Activity labels were manually identified by synchronizing the instrumented insole data with the video recordings. Turning events were informed by the definition provided by (30) in combination with observing the shoulder and pelvis rotation and foot orientation from the recorded videos, as well as observing the IMU signals for signs of pattern changes in foot acceleration and angular velocity. Stair ascent and descent started at the swing phase before the first foot came into contact with the stair tread and ended at the heel strike of the last foot to contact the floor/landing. Standing was identified as a period when the feet were in double support, and no other step was immediately initiated.

In Python, the sequential model from the Keras library (31) was used to develop artificial neural networks (ANNs) with four fully-connected dense layers with a decreasing number of hidden units for layer-wise feature compression (32). This ANN was used to identify five ambulatory activities: walking, standing, turning, stair ascent, and stair descent. The model’s first layer reshaped the input data for time-series analysis, followed by four dense layers with rectified linear unit activation functions for non-linear transformation. A time-distributed layer was used for dropout and regularization, followed by a flattening layer for vectorization. The final softmax layer performed the multi-class classification (Figure *1*1). Input data (i.e., raw pressure and IMU data) from the instrumented insoles were first scaled by subtracting the mean and dividing by the standard deviation and then reshaped into a vector. Activity classifications were made using a sliding window approach. Window sizes for each HAR model were selected heuristically from a predefined set of values (52, 100, 152, 200, or 252 frames, corresponding to 1.04 – 5.04 s of data) based on prior research (33), shown in Table *2*. The step size was always ¼ of the window size to balance overlap and computational efficiency (34). Windowed predictions were then reformatted as frames, and logic was applied to enforce real-world constraints (i.e., use knowledge of previous and next activity predictions to reevaluate short activities). To enforce standardization in the spatiotemporal analysis, periods identified as walking were indexed into 10-second segments.

**Figure 1.**
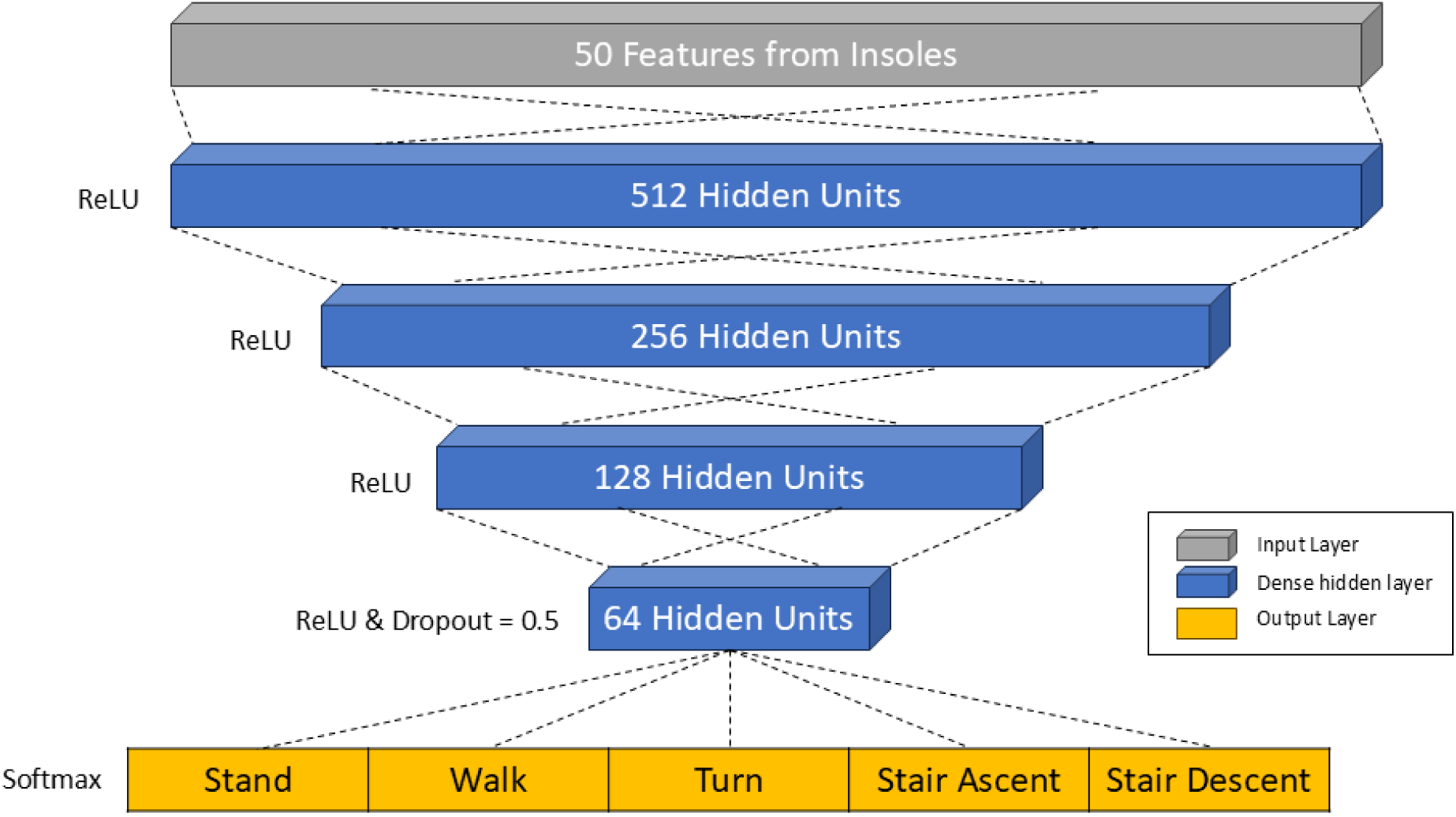
The architecture of the artificial neural network model used for gait activity recognition. The dense layers were fully-connected and had a diminishing number of hidden units for layer-wise feature compression. ReLU = rectified linear unit.

**Table 2.**
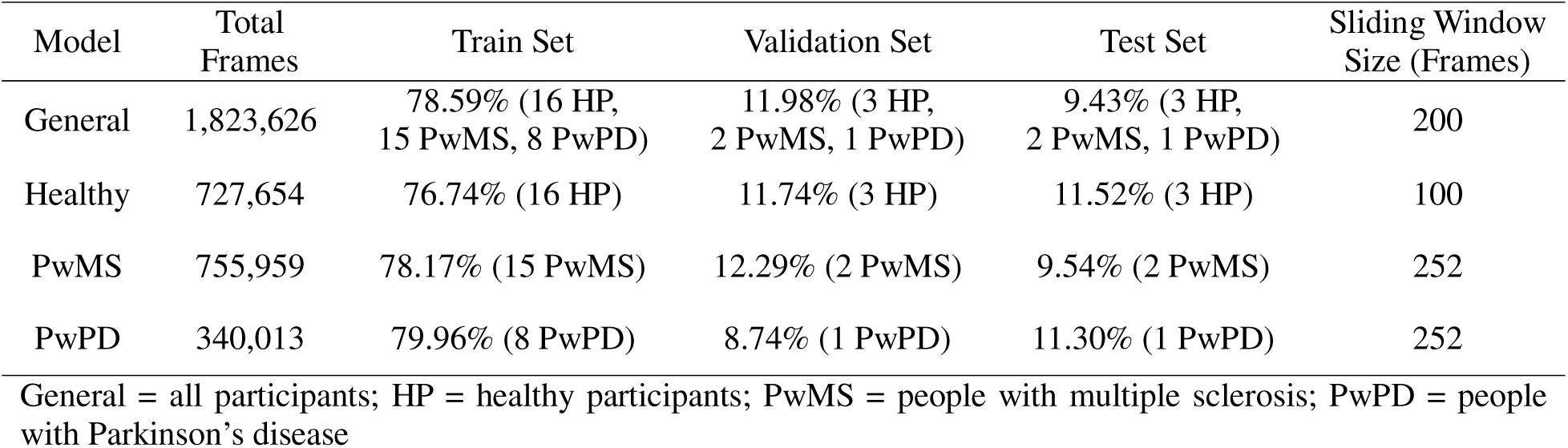
The data divisions and sliding window sizes for training and evaluating the artificial neural networks using hold-out testing.

Four HAR models were trained and evaluated: a General model (i.e., all participants included); a Healthy-specific model; an MS-specific model; and a PD-specific model. A hold-out testing approach was used to train and evaluate the models, where the randomly selected participants included in each train, validation, and test subsets are shown in Table *2*. Data from each participant were always kept in the same subset to avoid exposure during training. To accommodate for class imbalance (i.e., varying number of samples for each ambulatory activity), a class weight dictionary was computed using the training set and implemented during training using the “balanced” setting in the SciKit-Learn library. This setting implemented a heuristic method utilizing logistic regression to deal with rare events (35). The models were trained using a batch size of 24, maximum epochs of 500, and data were randomly shuffled before epochs to mitigate the learning of order effects (34). Loss was computed using the categorical cross-entropy loss function, where the gradients of the cost function were computed with respect to the parameters using backpropagation, where stochastic gradient descent updated the parameters to minimize loss. This was performed until the early stopping criterion was reached: training loss ceased to improve for five consecutive epochs. The model weights from the epoch with the lowest validation loss were saved for evaluation on the test set. Overall performance on the test set was evaluated using loss, accuracy, weighted averaged F1-score, and a confusion matrix; activity-specific performances were evaluated using precision, recall, and F1-score.

#### 2.3.2 Gait Detection

The gait detection algorithm used data from the pressure and IMU sensors to separate each gait cycle into four phases: heel strike (HES), foot on floor (FOF), heel rise (HER), and toe off (TOF), based on the definitions from Chatzaki and colleagues (15). Pressure data were pre-processed by grouping the 16 pressure sensors into three sections: heel (sensors 1-4), midfoot (sensors 5-7), and toes (sensors 9-16). IMU data were filtered using a 4th-order dual-pass Butterworth filter with a 6 Hz cut-off.

Gait detection began by independently identifying the middle of the swing phase (midSwing) using pressure and IMU data. midSwing was considered to be the location of the local valley in the pitch gyroscope signal (13) and the middle of a sustained period with pressure below 7% of the maximum observed pressure for that walking session (15). Logic was used to determine the final location of the midSwing using the information obtained from the pressure and IMU indices (i.e., relative location, relative values). Pre-swing was then discovered using methods described by Trojeniello et al. (13), defined as the region where the pitch gyroscope is ≥ 50% of the local peak while rising toward that peak. The location where all pressure sensor groups are minimized was considered the start of the TOF phase.(14)

After TOF, HES was independently determined by the pressure and IMU data. The IMU solution identified the valley in the forward accelerometer that occurs during the breaking phenomenon (13). The pressure solution identified the first frame where any pressure group surpassed 7% of the maximum observed value in the trial (15), which allowed any type of foot strike event (i.e., heel, midfoot, or toe strike). Logic was used to determine the final location of the HES event using the information obtained from the pressure and IMU indices. The HES event continued to be expressed until the vertical accelerometer and pitch gyroscope were within the empirically tested range of 0.9–1.1 g and -5–20 dps, respectively, which defined the FOF phase. The FOF phase continued while the vertical accelerometer and pitch gyroscope were within their respective range, the proper events proceeded (i.e., HES or FOF), and/or the midfoot pressure group was above the 7% threshold (15).

Finally, HER was found. Since this framework was designed to accommodate dysfunctional gait, the algorithm allows the person to transition between FOF and HER events when, for example, people lift their heel slightly during the stance phase but do not take a step forward (common in people using walkers who perform HES with their toes). The pre-swing indices found in the initial TOF calculations that have not been overwritten as another gait phase were automatically filled to be HER. Any remaining frames in the gait cycle are then evaluated for FOF; if they did not conform to the logic described above, they were labelled HER events. The gait detection algorithm is depicted in Figure 2.

**Figure 2.**
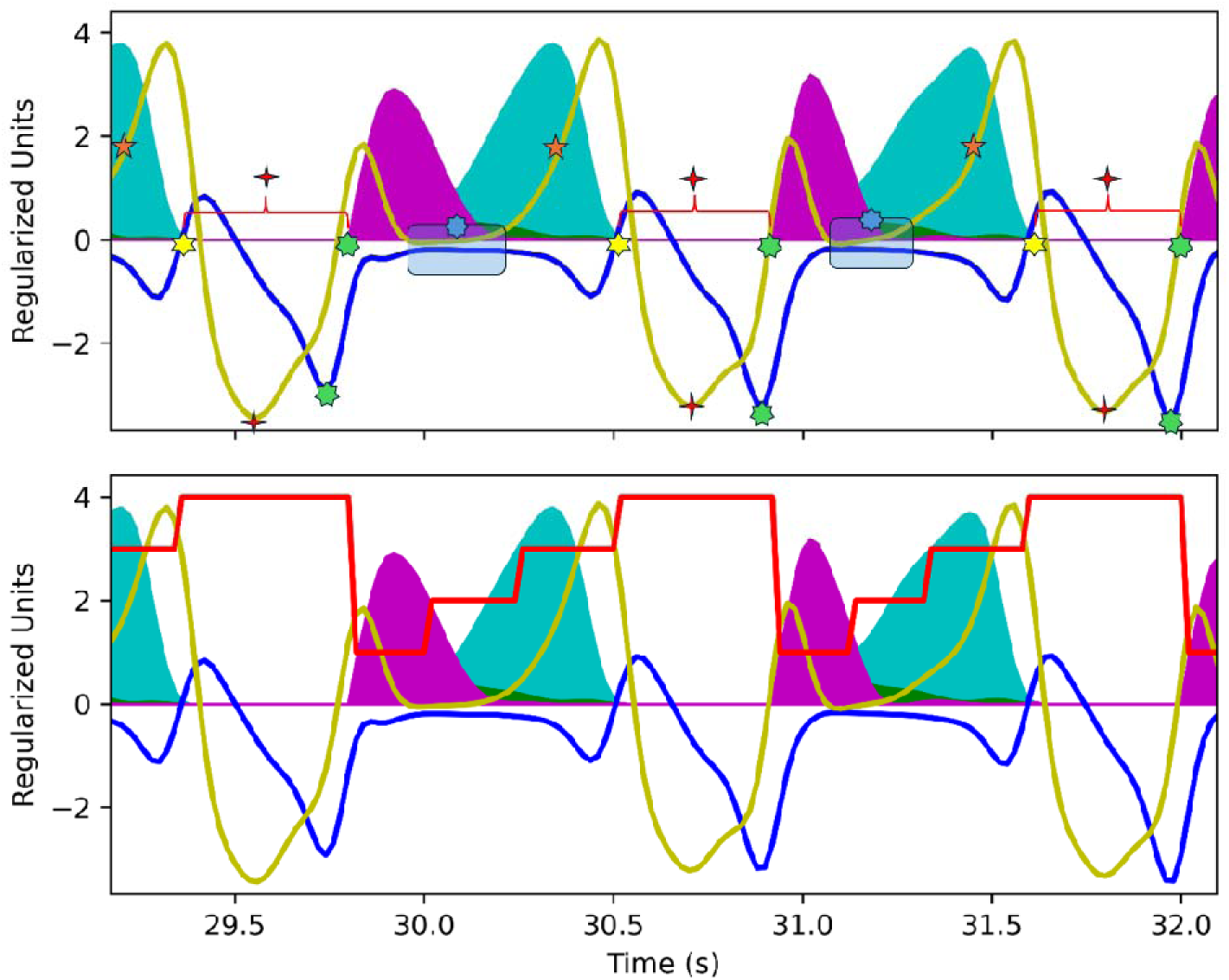
Gait Detection Algorithm. Depicted are the pressure and inertial measurement unit (IMU) data for the right insole for a healthy participant. Filled in curves illustrate pressure data: magenta = heel pressure, green = midfoot pressure, cyan = toe pressure. Lines illustrate the forward accelerometer (blue), pitch gyroscope (yellow), and gait phases (red), where one = heel strike, two = foot on floor, three = heel rise, and four = toe off. The upper figure depicts the logic used to determine gait phases. The order of operations can be followed using the stars coloured using rainbow convention and the number of star points. First (4-point red star), midswing is identified using pressure and IMU data. Second (orange 5-point star), pre-swing is found using the gyroscope data. Third (yellow 6-point star), toe off is found by minimizing the pressure after pre-swing. Fourth (green 7-point star), heel strike is found using the IMU and pressure data. Fifth (blue 8-point star) foot on floor is found using an apriori IMU range. Sixth, all points not identified as foot on floor are considered heel rise.

#### 2.3.3 Sensor Fusion

To remove the bias in the gyroscope signal, the median value for all three axes was calculated using all of the standing activities identified through HAR and subtracted from the signals. Then, a Madgwick filter with a zero-velocity update was applied to the raw IMU data (gyroscope bias-corrected) to obtain the linear acceleration of each foot in global space (36). The global linear acceleration was then integrated to obtain velocity and was drift-corrected using the stationary period within the stance phase (37). The corrected velocity was then integrated to obtain the foot’s position. The gain value used for this analysis was determined by minimizing the root mean squared error between the position calculated through the IF and the MoCap systems.

### 2.4 Motion Capture (MoCap) Data

For all laboratory-based movements, video camera data were processed in Theia 3D (Theia Markerless Inc., Canada) to obtain whole-body kinematics. Gait events were calculated by importing the kinematic and kinetic data into Visual 3D (V5, HAS-Motion, Canada) and using their automatic gait detection pipeline. These events were imported into Vicon Nexus 2.12 (Vicon, UK), where the events were visually verified and adjusted as necessary. Spatial, temporal, spatiotemporal and asymmetry variables were calculated using ProCalc (Vicon, UK) and custom algorithms in Matlab 2018b (MathWorks, USA).

### 2.5 Gait Metrics Calculated by the Insole Framework (IF)

Gait analyses in this investigation focused on relatively straight walking between turns, standing, or stair ascent/descent activities. When walking trials were longer than 30 seconds (i.e., 125- and 500-metre indoors and 500-metre outdoors), these walks were separated into 10-second segments. In cases where a period of walking was interrupted by another ambulatory activity and the walking duration was greater than 10 seconds but less than 20 seconds (or any other base ten value), 10-second segments were taken as the middle portion of the walk. For example, if a participant turned after 16 seconds of straight walking, the first and last three seconds would be discarded, and the middle 10 seconds would be analyzed as described below.

Using the identified gait events, spatiotemporal metrics were calculated and grouped into four categories: core, pace, percentage, and asymmetry. A summary of spatiotemporal metric descriptions and calculations is presented in the Supplementary Material.

Core spatiotemporal metrics were metrics from which all other categories’ metrics were calculated. Within the same foot, stride time was the time between HES events; stance time was the time between HES and TOF events; swing time was the time between TOF and HES events; and stride length was the arclength distance between HES events. Between feet, step time was the time between HES events of opposing feet; single support time was the time when only one foot was in the stance phase; and double support time was the time when both feet were in the stance phase (initial and terminal double support were combined).

Pace metrics describe how fast the participant was moving. Cadence was calculated by multiplying the step time by 60 seconds to obtain steps per minute (steps/min). Stride velocity was the stride length divided by the stride time to obtain metres per second (m/s).

Percentage metrics were temporal-based core metrics normalized to a percent of stride time as shown in Equation 1:

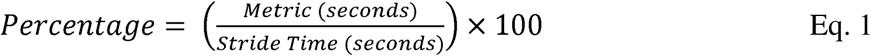

Asymmetry metrics were core metrics expressed as a percent difference between sides of the body. Asymmetry was calculated by first rounding the metrics to two decimal places and then following Equation 2 (38).

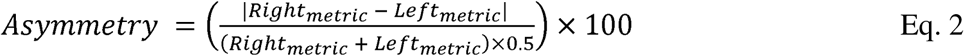

### 2.5 Statistical Analysis

When appropriate, MoCap and insole data were aligned using the HES events that occurred at the force plate. The events were manually identified for each foot, and the average offset between feet was used to update the timestamp of the MoCap system. If an HES event did not align with the rising edge of the vertical force values (e.g., an assistive device contacted the force plate first; HES happened on a force plate that they were already standing on), two HES events per foot located anywhere in the trial were manually identified and used to find the average temporal offset to update the MoCap time stamp.

Because the MoCap system and the insoles were a random sample of their respective system (i.e., MoCap can have many calibrations and camera placements; insoles come in different sizes and variations in manufacturing), and because future use will only use the measurements of a single rater (39), two-way random effects with *consistency* and *single-rater* intraclass correlations (ICC_2,1_) were calculated to assess the reliability of the IF for calculating spatial, temporal, and spatiotemporal metrics (39) for all overground walks in the laboratory. Bland-Altman Limits of Agreement (LoA) were also calculated for each gait metric to assess the agreement between the two systems. ICC_2,1_ and LoA were calculated separately for the healthy, PwMS, and PwPD populations to assess any differences between populations.

One-way analysis of variance tests (ANOVAs) were run to identify significant differences between populations to evaluate whether both systems could identify the same significance trends between populations. When parametric assumptions were violated (i.e., through significant Levene’s tests and/or Shapiro-Wilk), Kruskal-Wallis (KW) tests were performed. To avoid type one errors, a Bonferroni correction was performed for the group-wise comparisons (i.e., 0.05/19 tests = significance at *p* < 0.0026) and post-hoc tests (i.e., 0.05/3 tests = significance at *p* < 0.0167).

## 3.0 Results

For all ICC results, the reliability of the IF was interpreted as excellent (> 0.90), good (0.75-0.90), moderate (0.50-0.75), or poor (< 0.50) as per Koo and Li (39).

### 3.1 Human Activity Recognition (HAR)

The overall classification performances of each ANN are presented in Table *3*, and the activity-specific performances are presented in Table 4. The confusion matrices for predictions on the test sets for all ANNs are presented in Figure 3.

**Figure 3.**
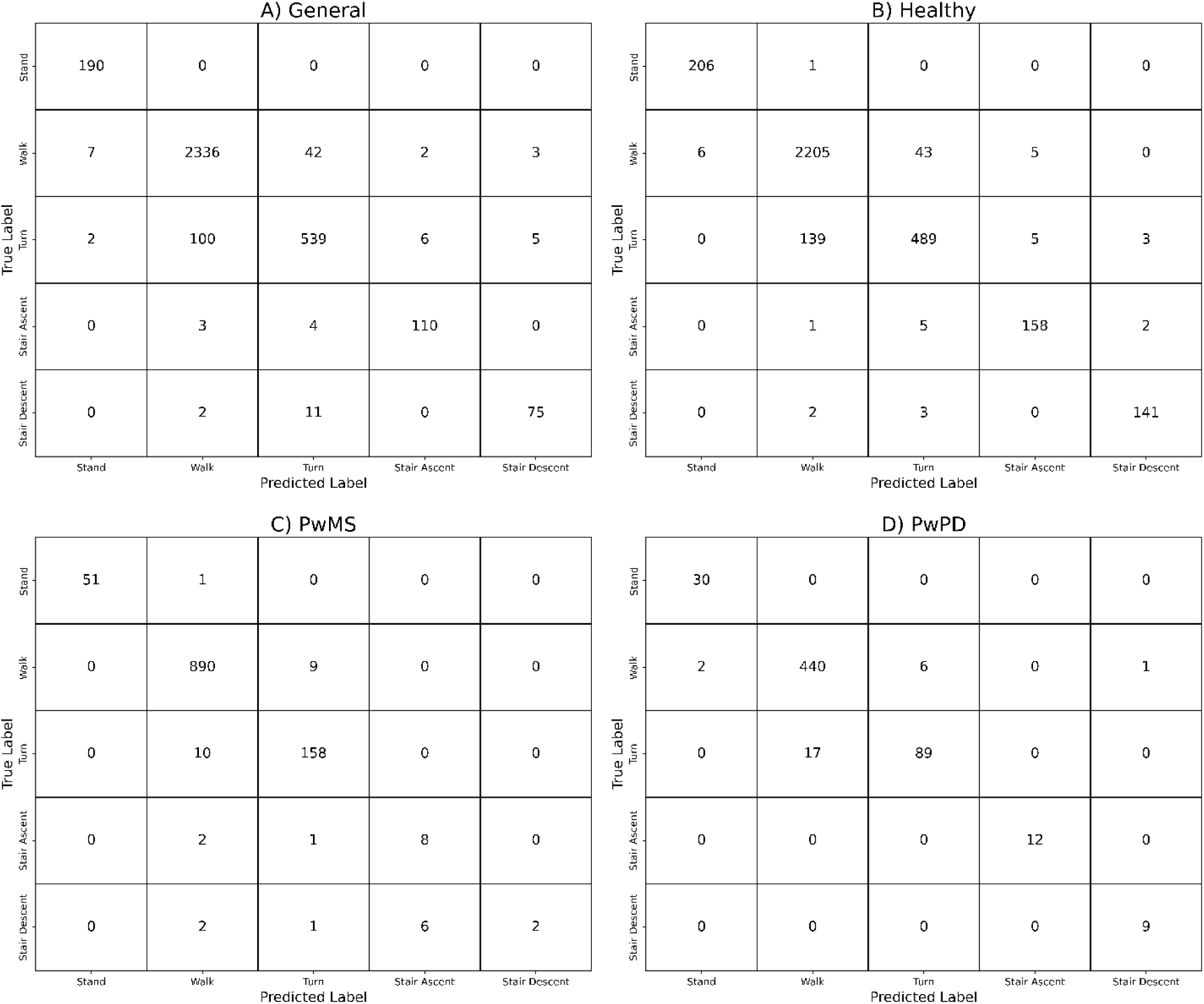
Confusion matrices for the predictions made by the following artificial neural networks: A) General (all participants); B) Healthy; C) People with multiple sclerosis (PwMS); and D) People with Parkinson’s disease (PwPD).

**Table 3.**
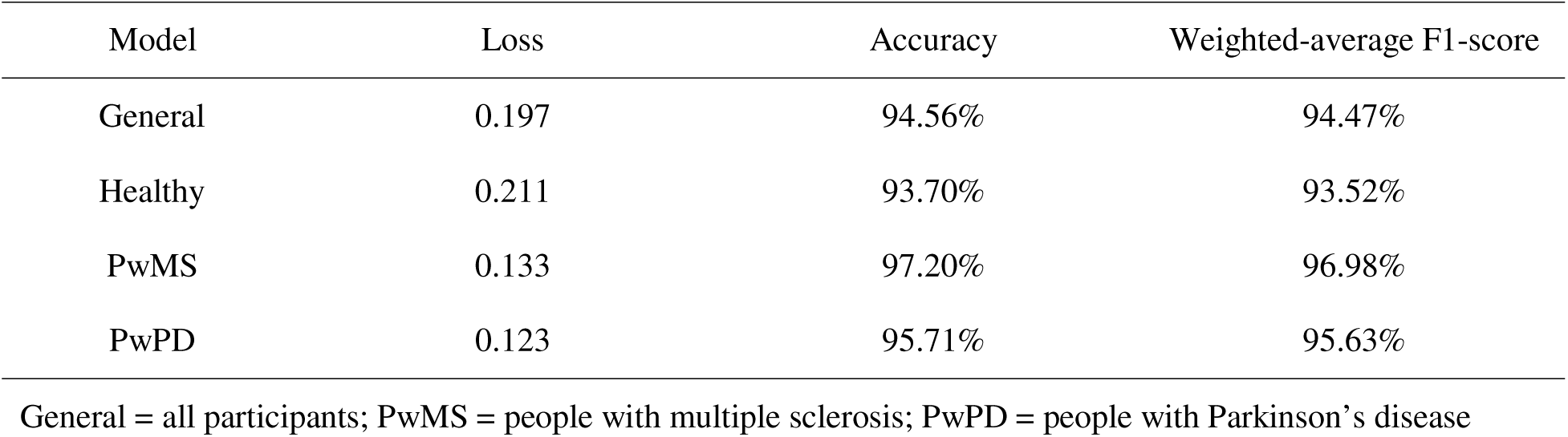
Classification performance for each gait activity recognition model.

**Table 4.** Activity-specific classification performances for each artificial neural network.

| Model | Performance Metric | Stand | Walk | Turn | Stair Ascent | Stair Descent |
| --- | --- | --- | --- | --- | --- | --- |
| General | Precision | 0.955 | 0.957 | 0.904 | 0.932 | 0.904 |
|  | Recall | 1.000 | 0.977 | 0.827 | 0.940 | 0.852 |
|  | F1-Score | 0.977 | 0.967 | 0.864 | 0.936 | 0.877 |
| Healthy | Precision | 0.972 | 0.939 | 0.906 | 0.940 | 0.966 |
|  | Recall | 0.995 | 0.976 | 0.769 | 0.952 | 0.966 |
|  | F1-Score | 0.983 | 0.957 | 0.832 | 0.946 | 0.966 |
| PwMS | Precision | 1.000 | 0.983 | 0.935 | 0.571 | 1.000 |
|  | Recall | 0.981 | 0.990 | 0.940 | 0.727 | 0.182 |
|  | F1-Score | 0.990 | 0.987 | 0.938 | 0.640 | 0.308 |
| PwPD | Precision | 0.938 | 0.963 | 0.937 | 1.000 | 0.900 |
|  | Recall | 1.000 | 0.980 | 0.840 | 1.000 | 1.000 |
|  | F1-Score | 0.968 | 0.971 | 0.886 | 1.000 | 0.947 |

General = all participants; PwMS = people with multiple sclerosis; PwPD = people with Parkinson’s disease

### 3.2 Reliability and Agreement

Nineteen spatiotemporal gait metrics were compared between the MoCap system and the IF. The average ICC_2,1_ value for core, pace, percentage, and asymmetry metrics were 0.938 ±0.065, 0.981 ±0.007, 0.664 ±0.045, and 0.553 ±0.226, respectively, for healthy participants; 0.957 ±0.051, 0.981 ±0.021, 0.854 ±0.011, and 0.876 ±0.082, respectively, for PwMS; and 0.965 ±0.0.045, 0.991 ±0.002, 0.848 ±0.031, 0.834 ±0.092, respectively for PwPD. Degrees of freedom for the healthy population are 21, 21; 17, 17 for the PwMS; and 9, 9 for the PwPD. ICC_2,1_ values for each metric are presented in Table 5. LoA results were similar between populations, and overall results were considered acceptable, with temporal metrics having a bias generally within the limitations of both systems’ sampling frequencies (i.e., 0.02 seconds; 50 Hz) and stride length having a bias of less than 2%. All LoA results are presented in Table 6 and visually depicted in the Supplementary Material.

**Table 5.** ICC_2,1_ results comparing the motion capture system and Insole Framework.

| Category | Metric | Population | ICC<sub>2,1</sub> [95% CI] | F-value | Interpretation |
| --- | --- | --- | --- | --- | --- |
| Core | Stride Time (s) | HP | 0.998 [1.00, 1.00] | 1111 | Excellent |
|  |  | PwMS | 1.000 [1.00, 1.00] | 8615 | Excellent |
|  |  | PwPD | 0.998 [0.99, 1.00] | 1229 | Excellent |
|  | Stance Time (s) | HP | 0.974 [0.94, 0.99] | 74.85 | Excellent |
|  |  | PwMS | 0.985 [0.96, 1.00] | 298.6 | Excellent |
|  |  | PwPD | 0.983 [0.94, 1.00] | 121.3 | Excellent |
|  | Swing Time (s) | HP | 0.884 [0.75, 0.95] | 20.52 | Good |
|  |  | PwMS | 0.894 [0.75, 0.96] | 22.08 | Good |
|  |  | PwPD | 0.954 [0.83, 0.99] | 62.42 | Excellent |
|  | Single Support Time (s) | HP | 0.929 [0.84, 0.97] | 27.26 | Excellent |
|  |  | PwMS | 0.882 [0.72, 0.96] | 18.53 | Good |
|  |  | PwPD | 0.960 [0.85, 0.99] | 58.50 | Excellent |
|  | Double Support Time (s) | HP | 0.824 [0.62, 0.93] | 10.38 | Good |
|  |  | PwMS | 0.948 [0.87, 0.98] | 37.51 | Excellent |
|  |  | PwPD | 0.870 [0.57, 0.97] | 14.37 | Good |
|  | Stride Length (m) | HP | 0.965 [0.95, 0.99] | 89.49 | Excellent |
|  |  | PwMS | 0.993 [0.98, 1.00] | 307.3 | Excellent |
|  |  | PwPD | 0.991 [0.97, 1.00] | 348.3 | Excellent |
|  | Step Time (s) | HP | 0.997 [0.99, 1.00] | 586.6 | Excellent |
|  |  | PwMS | 1.000 [1.00, 1.00] | 6204 | Excellent |
|  |  | PwPD | 0.997 [0.99, 1.00] | 757.5 | Excellent |
| Pace | Cadence (steps/min) | HP | 1.000 [0.99, 1.00] | 494.1 | Excellent |
|  |  | PwMS | 0.966 [0.91, 0.99] | 58.26 | Excellent |
|  |  | PwPD | 0.990 [0.96, 1.00] | 193.7 | Excellent |
|  | Stride Velocity (m/s) | HP | 0.976 [0.94, 0.99] | 95.73 | Excellent |
|  |  | PwMS | 0.996 [0.99, 1.00] | 506.8 | Excellent |
|  |  | PwPD | 0.993 [0.97, 1.00] | 321.2 | Excellent |
| Percentages | Stance Percent (%) | HP | 0.669 [0.39, 0.85] | 5.370 | Moderate |
|  |  | PwMS | 0.852 [0.67, 0.94] | 20.95 | Good |
|  |  | PwPD | 0.872 [0.58, 0.97] | 15.04 | Good |
|  | Swing Percent (%) | HP | 0.669 [0.39, 0.85] | 5.370 | Moderate |
|  |  | PwMS | 0.852 [0.67, 0.94] | 20.95 | Good |
|  |  | PwPD | 0.872 [0.58, 0.97] | 15.04 | Good |
|  | Single Support Percent (%) | HP | 0.766 [0.52, 0.90] | 7.754 | Good |
|  |  | PwMS | 0.868 [0.70, 0.95] | 23.72 | Good |
|  |  | PwPD | 0.839 [0.49, 0.96] | 12.38 | Good |
|  | Double Support Percent (%) | HP | 0.720 [0.44, 0.87] | 6.193 | Moderate |
|  |  | PwMS | 0.841 [0.63, 0.94] | 11.66 | Good |
|  |  | PwPD | 0.807 [0.40, 0.95] | 9.387 | Good |
| Asymmetry | Stride Time Asymmetry (%) | HP | 0.838 [0.65, 0.93] | 11.34 | Good |
|  |  | PwMS | 0.960 [0.90, 0.98] | 48.42 | Excellent |
|  |  | PwPD | 0.700 [0.17, 0.92] | 5.674 | Moderate |
|  | Stance Time Asymmetry (%) | HP | 0.702 [0.41, 0.86] | 5.710 | Moderate |
|  |  | PwMS | 0.866 [0.98, 0.95] | 13.88 | Good |
|  |  | PwPD | 0.870 [0.56, 0.97] | 14.44 | Good |
|  | Swing Time Asymmetry (%) | HP | 0.506 [0.12, 0.76] | 3.048 | Moderate |
|  |  | PwMS | 0.864 [0.67, 0.95] | 13.75 | Good |
|  |  | PwPD | 0.973 [0.89, 0.99] | 72.20 | Excellent |
|  | Stride Length Asymmetry (%) | HP | 0.616 [0.27, 0.82] | 4.205 | Moderate |
|  |  | PwMS | 0.731 [0.41, 0.89] | 6.435 | Moderate |
|  |  | PwPD | 0.841 [0.48, 0.96] | 11.57 | Good |
|  | Single Support Time Asymmetry (%) | HP | 0.177 [-0.25, 0.55] | 1.432 | Poor |
|  |  | PwMS | 0.893 [0.74, 0.96] | 17.70 | Good |
|  |  | PwPD | 0.847 [0.50, 0.96] | 12.10 | Good |
|  | Double Support Time Asymmetry (%) | HP | 0.479 [0.08, 0.75] | 2.842 | Poor |
|  |  | PwMS | 0.945 [0.86, 0.98] | 35.46 | Excellent |
|  |  | PwPD | 0.773 [0.32, 0.94] | 7.807 | Good |

Note: HP = healthy participants, PwMS = people with multiple sclerosis, PwPD = people with Parkinson's disease, s = seconds, m = metres, min = minutes, m/s = metres/second, and % = percentage. Percentage variables are the percent of stride time, and asymmetry variables are the percent difference between sides (Eq 2).

**Table 6.**
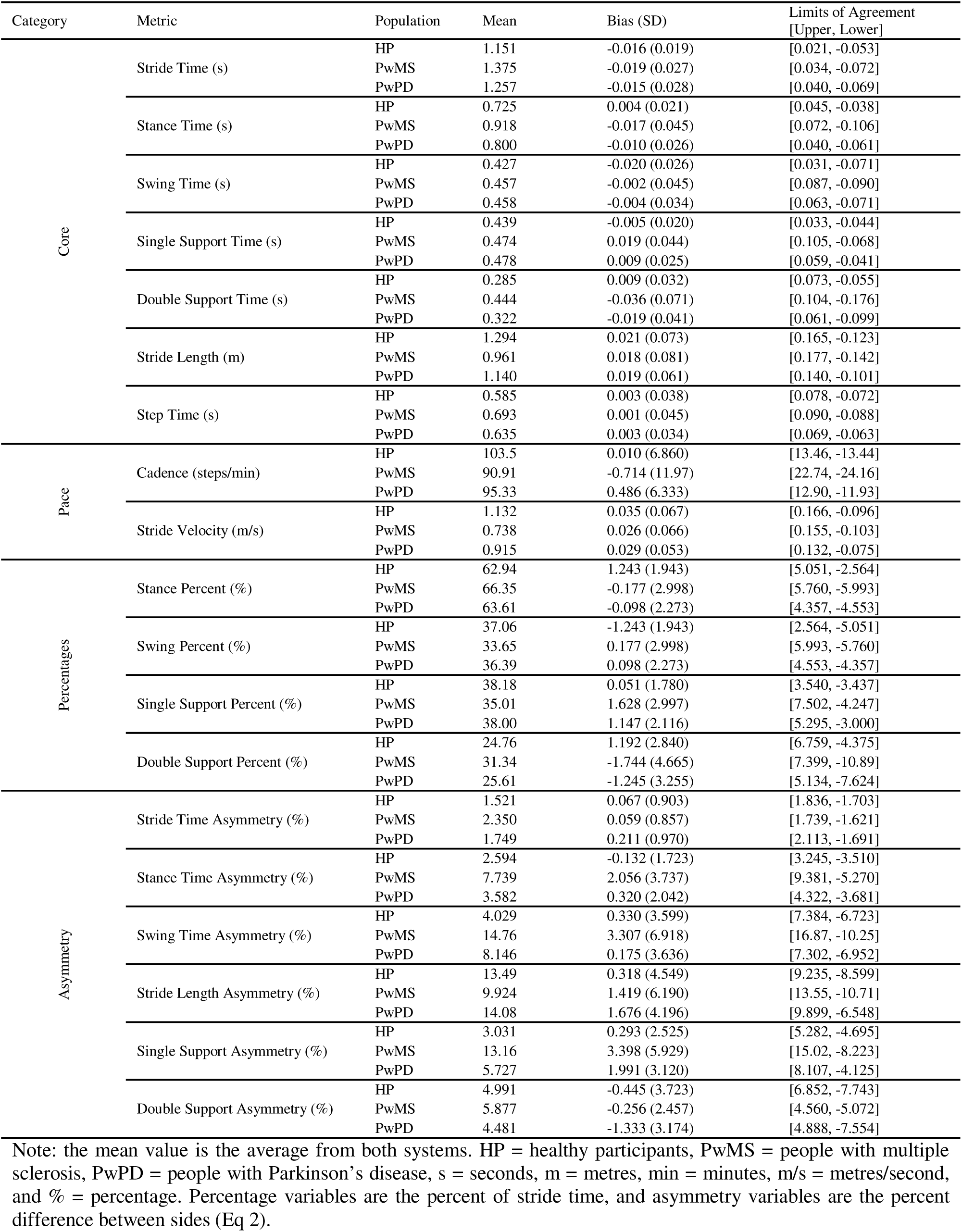
Bland Altman Limits of Agreement Results comparing the motion capture system and Insole Framework.

### 3.3 Significance Testing

Nineteen spatiotemporal metrics were independently assessed for significant differences between populations (*p* < 0.0026) using the IF and the MoCap system. All but one metric (i.e., stride length asymmetry) had the same significance interpretation between systems. For those spatiotemporal metrics that had significant differences between populations (N = 10), the systems disagreed on three metrics during post-hoc testing where the MoCap system found no difference between PwMS and PwPD during stance, swing, and double support percent, but the IF did (*p* < 0.0167). Significance testing results are presented in Table 7.

**Table 7.**
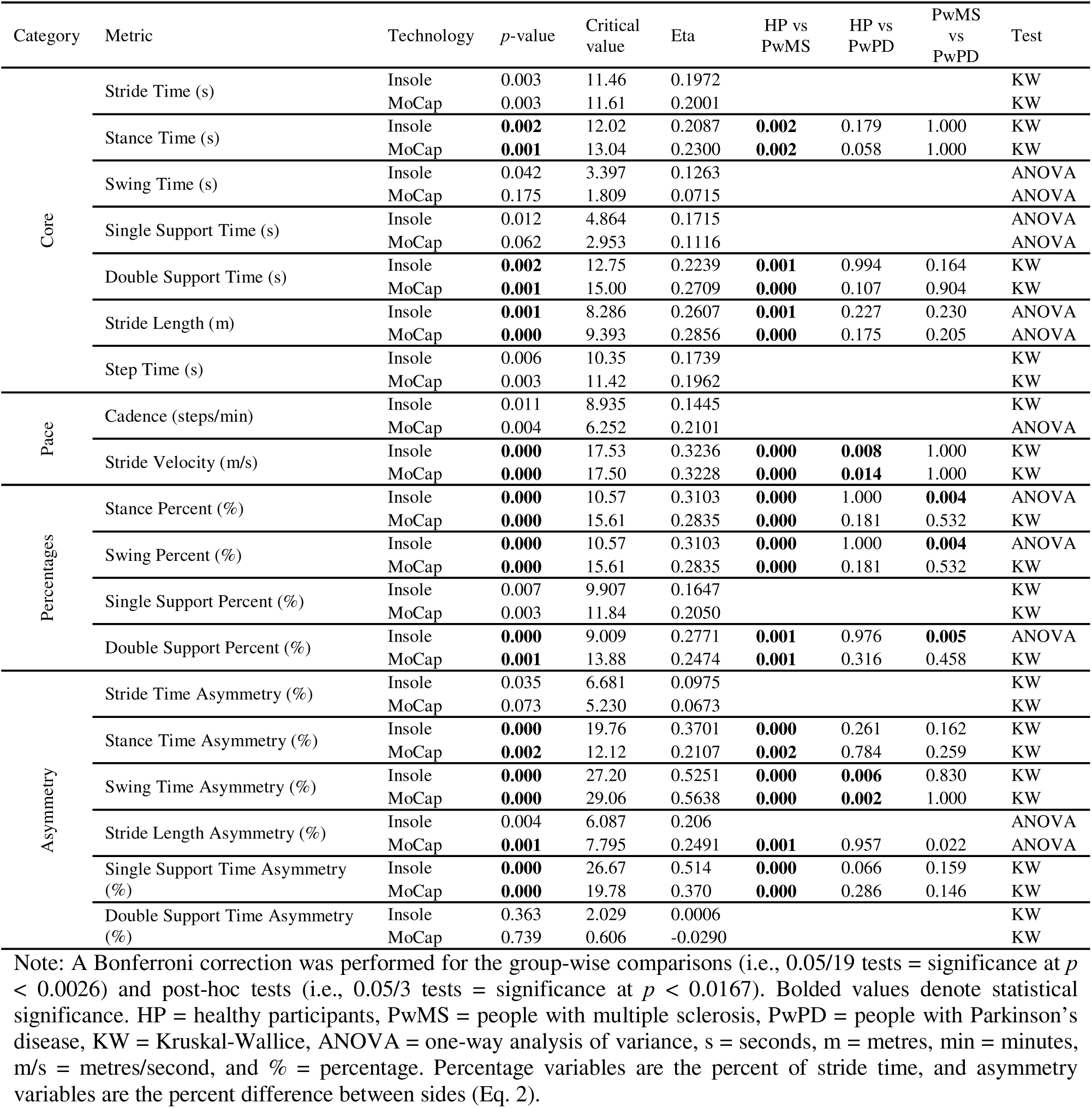
Statistical Comparisons Between Populations.

## 4.0 Discussion

Evaluating walking quality is a key method for understanding disease progression in people who have neurological disorders (40). However, the current standard of care focuses on gross walking ability, such as walking speed and fall history, rather than spatiotemporal gait metrics, mainly due to the barriers to accessing the technology needed and the frequency with which these metrics need to be collected to be valuable for longitudinal evaluations and proactive decision-making. Therefore, we propose a method that uses instrumented shoe insoles to collect the data needed to unobtrusively monitor the gait quality of individuals outside of a laboratory environment. The current paper validates an IF that uses a multi-layered approach to automatically identify ambulatory activities (i.e., walking, standing, turning, stair ascent, and stair descent), perform gait detection, and calculate reliable spatiotemporal gait metrics on standardized 10-second segments of walking.

Our HAR models showed strong performance (i.e., the General model had an accuracy of 94.56% and weighted-averaged F1-score of 94.47%), which is comparable to previous literature using instrumented shoe insoles to detect similar types and numbers of activities (21,23,24). However, the HAR models developed with data solely from individuals with neurological dysfunction (i.e., MS and PD) led to improved classification performances (accuracies ranged from 95.71 - 97.20%) compared to the General model. This was likely because the variability in gait patterns within a population was equal to or lesser than the General model, improving the ANN’s ability to correctly relate the raw insole data to gait activities. These findings should motivate the use of population-specific modelling for HAR. Consistent with previous literature (23), our algorithms generally had a lower precision for stair ascent and descent compared to other activities, especially in the PwMS-specific model, which often incorrectly predicted stair ascent when the true label was stair descent. It is possible that similarities in the kinematics and kinetics of the first and last stair tread of stair ascent versus descent may have contributed to the poorer performance in these activities. While classification performances for the stair activities were stronger in the other models, the larger variance in gait parameters amongst PwMS compared to healthy controls (41) may have posed a greater challenge to recognizing their stair activities: more training examples are warranted. The results of the present work suggest that classifying ambulatory activities for PwMS (and potentially other disorders affecting gait) may be further strengthened by developing HAR models specific to gait phenotypes (e.g., ataxic, spastic, hemiplegic, etc.).

Compared to the gold-standard MoCap system, the IF was able to calculate all core and pace spatiotemporal gait metrics with good to excellent reliability (≥ 0.824) regardless of neurological status. All spatiotemporal metrics had moderate to excellent reliability for PwMS (≥ 0.731) and PwPD (≥ 0.700), while HP had moderate to good reliability for percent metrics (0.669-0.766) and poor to good reliability for asymmetry metrics (0.177-0.838). On average, across all metrics, PwMS had the highest ICC_2,1_ values (0.912), followed by PwPD (0.902); HP had the lowest average ICC_2,1_ values (0.773). LoA results indicate that bias values for core temporal metrics across all populations are within the limitations of the sampling frequencies (i.e., 0.02 seconds) except for double support time in PwMS (0.036 seconds). On average, PwMS had the highest bias values, followed by PwPD, and HP had the lowest biases.

Significance testing using one-way ANOVAs and Kruskal-Wallis tests indicated that similar statistical findings would be discovered if analyzing the same population using either the IF or the MoCap system. Of the 19 spatiotemporal metrics assessed, the systems disagreed on one population-level test (i.e., stride length asymmetry) and three post-hoc comparisons (i.e., PwMS vs. PwPD for stance, swing, and double support percent). Further, across all tests, η^2^ values – which represent the proportion of variance explained by group differences – were relatively consistent between technologies; the average absolute η^2^ difference across all tests was 0.046. This consistency suggests that, regardless of the system used, a similar amount of variance was explained for the captured group-based differences.

Overall, the presented temporal results are comparable to previous research that reported ICC (42–45) and LoA values (16–18,44) when comparing instrumented insoles to a gold-standard system. Stride length results in the current study are similar to what has been previously reported with foot-mounted IMUs (37,44,45) and instrumented shoe insoles (16,18), reaffirming that using a fusion algorithm (i.e., Madgwick Filter (36) in the current study) with a zero-velocity update is appropriate to use in healthy and dysfunctional gait. Percentage and asymmetry gait metrics had the lowest reliability in the healthy population, which is similar to findings from a review by Kobsar et al. (46), who found that throughout the literature, asymmetry and variability metrics have poor reliability in healthy populations. For PwMS and PwPD, moderate to excellent reliability was found for asymmetry metrics; the stronger reliability is likely due to the increased between-subject variability in gait quality amongst the PwMS and PwPD populations, favouring ICC calculations compared to the more homogenous HP (39).

The presented work has limitations. Due to distance constraints, the laboratory work was limited to approximately six metres of walking. This meant that participants may not have achieved steady-state walking, which is the expected behaviour when walking in the wild. Although collected, treadmill walking data was not used for this investigation; however, future work will focus on these data to develop gait stability analyses. Moreover, the trained HAR algorithm is limited in its scope for detecting ambulatory activities. People perform many more activities during their daily lives; however, the expected use case for the presented framework is during purposeful walking, which would limit the number of arbitrary activities performed during the walking assessment. Pressure and IMU data were used for the gait detection algorithms, but metrics were not calculated using these data since they could not be directly compared to the MoCap system. Future work aimed at identifying useful metrics that can be obtained from these sensors, as well as the reliability of these metrics, is suggested. While the current work demonstrates a valid framework for collecting and analyzing human gait in HP, PwMS, and PwPD, the metrics reported here and throughout the literature are not always interpretable for the average clinician who is not a gait expert. Future work should focus on developing accessible methods and/or platforms for presenting relevant metrics to clinicians and patients for improved monitoring of gait quality over time, and to help inform treatment decisions.

## 5.0 Conclusion

Presented is a framework to analyze human gait using instrumented shoe insoles. Our IF performs activity recognition to identify ambulatory activities, performs gait detection by using both pressure and IMU data, standardizes the analysis into 10-second segments, and reliably calculates spatiotemporal metrics in HP, PwMS, and PwPD. The presented results instill confidence that our IF can calculate traditional spatiotemporal gait metrics in healthy and dysfunctional gait.

## Supporting information

Supplemental Bland Altman Plots

Supplemental Variable Descriptions

## Data Availability

Spatiotemporal data from the insoles and motion capture system are available as an excel file in the Supporting Information files.

## 6.0 Acknowledgements

## Author contributions

conceptualization: MPM, RBG, MSF, HM, GB. Data curation: MPM, AMO. Formal analysis: MPM, AMO, VCHC. Funding Acquisition: MPM, RBG, MSF. Investigation: MPM, AMO. Methodology: MPM, AMO, VCHC. Project Administration: RBG. Resources: RGB, MSF, GB, HM, DG, TM. Software: MPM, AMO, VCHC. Supervision: RBG. Validation: MPM. Visualization: MPM, VCHC. Writing – original draft: MPM. Writing – review: all authors.

