## Supplemental Bland Altman Plots for "Validation of an instrumented shoe insole framework for analyzing spatiotemporal gait metrics in healthy and neurodegenerative populations"

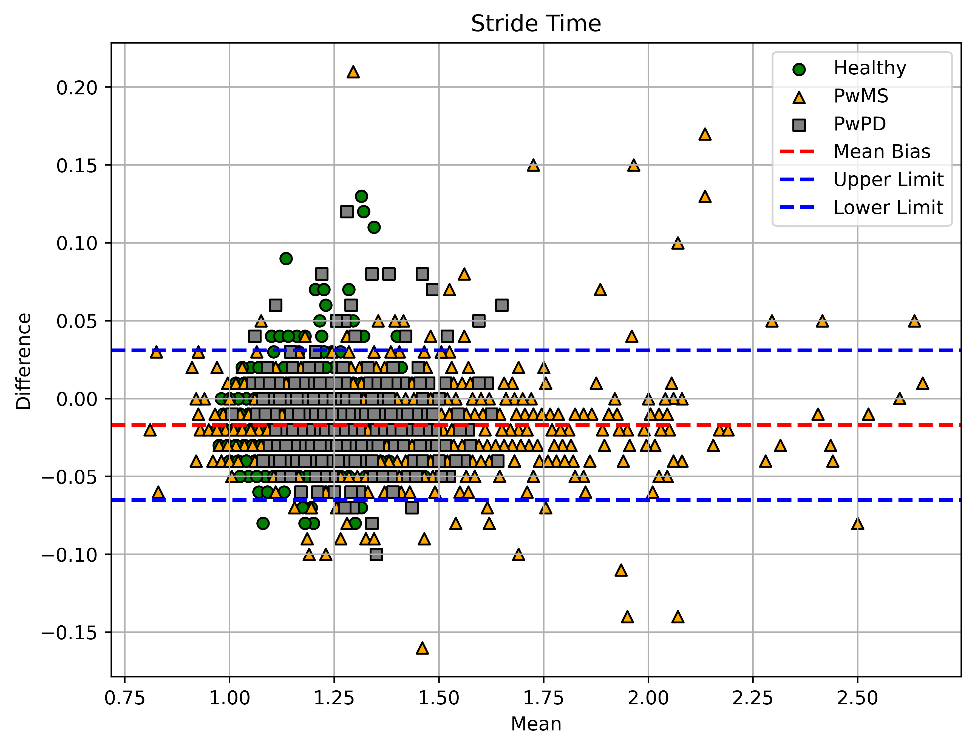

### Core Metrics

Figure 1 Stride Time Bland Altman Limits of Agreement Plot. PwMS = people with multiple sclerosis, PwPD = people with Parkinson’s disease. Bias and upper and lower limits were calculated using all participants.

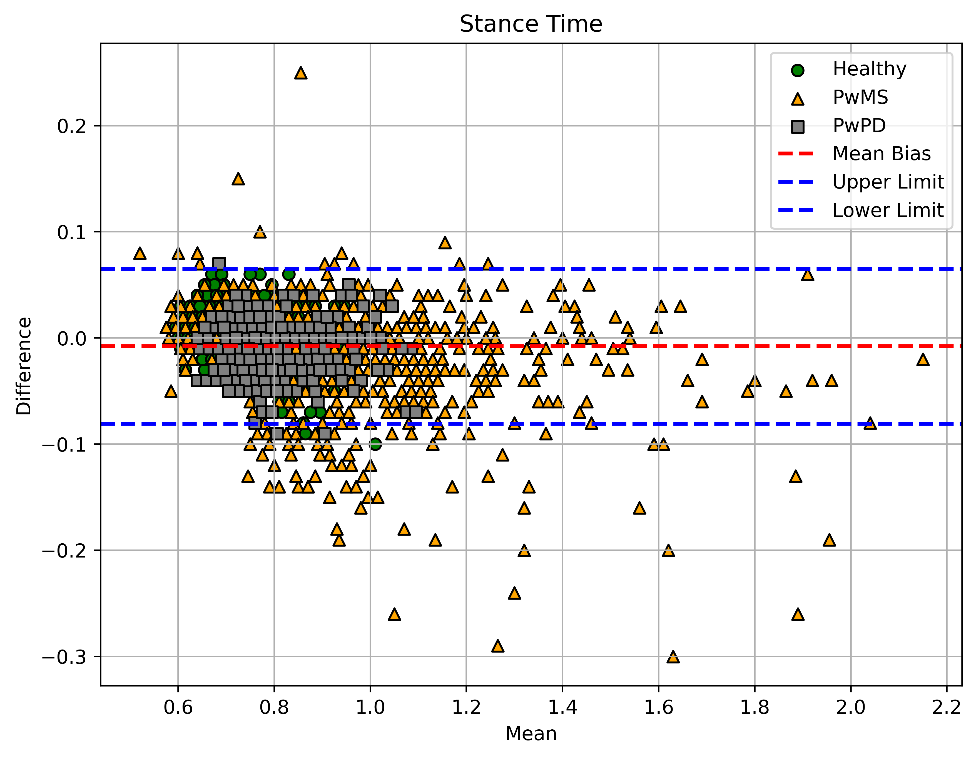

Figure 2 Stance Time Bland Altman Limits of Agreement Plot. PwMS = people with multiple sclerosis, PwPD = people with Parkinson’s disease. Bias and upper and lower limits were calculated using all participants.

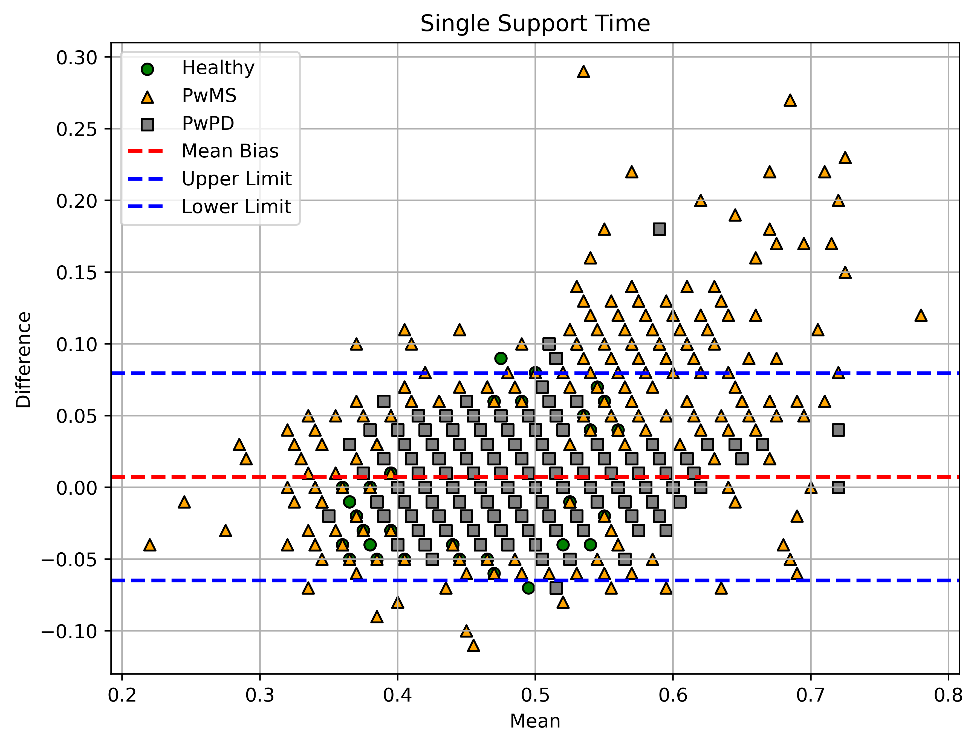

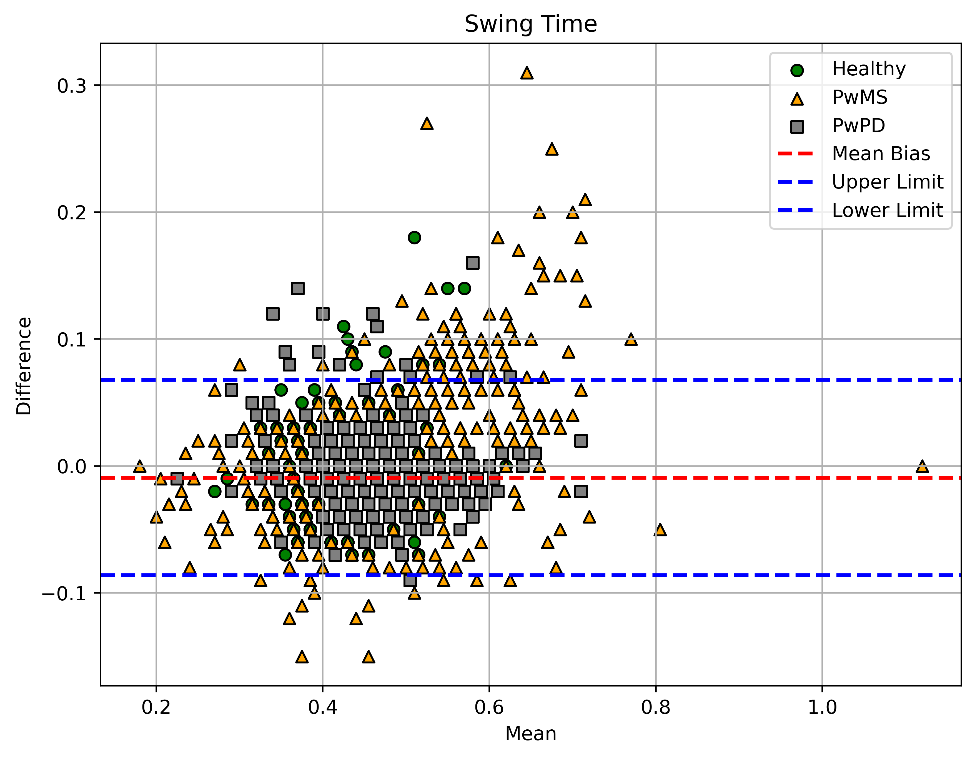

Figure 3 Swing Time Bland Altman Limits of Agreement Plot. PwMS = people with multiple sclerosis, PwPD = people with Parkinson’s disease. Bias and upper and lower limits were calculated using all participants.

Figure 4 Single Support Time Bland Altman Limits of Agreement Plot. PwMS = people with multiple sclerosis, PwPD = people with Parkinson’s disease. Bias and upper and lower limits were calculated using all participants.

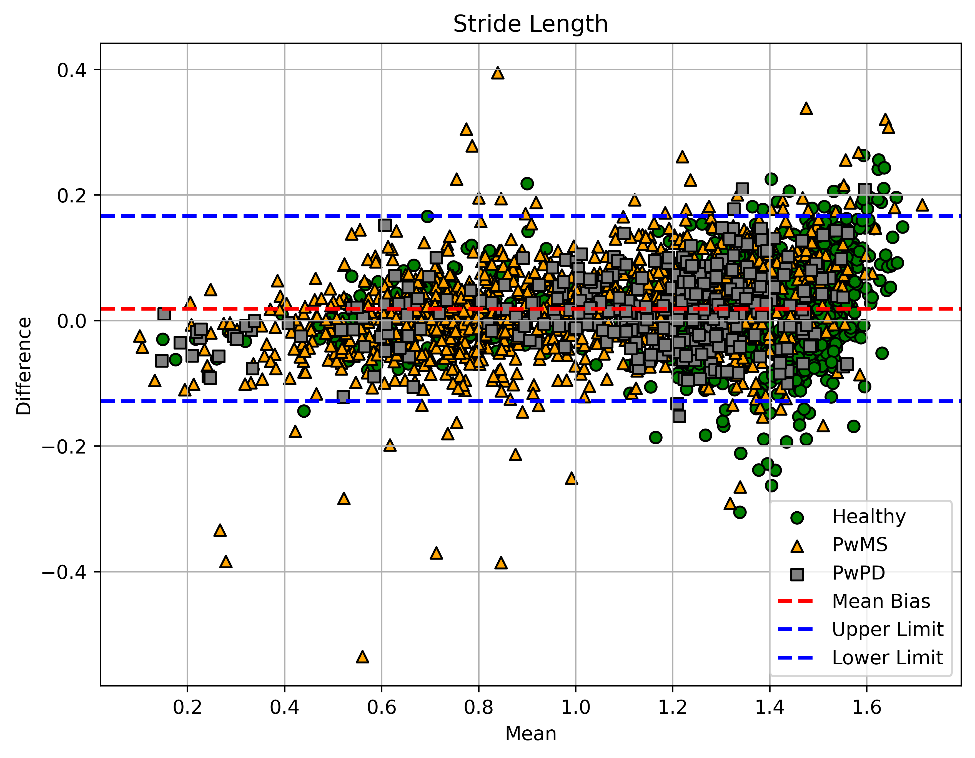

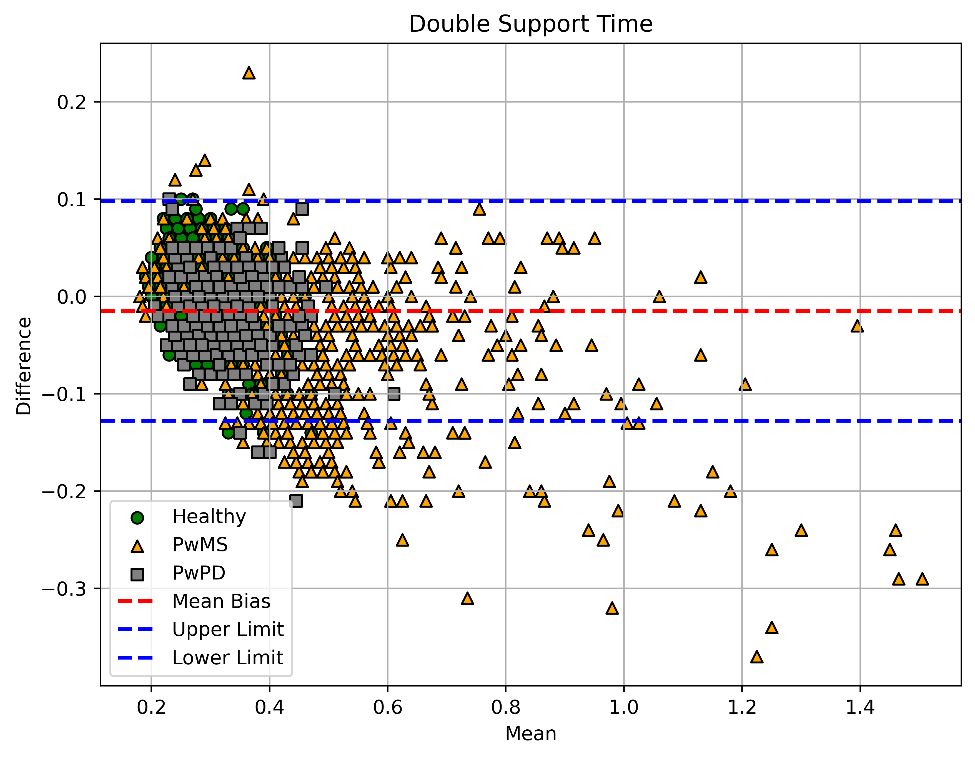

Figure 5 Double Support Time Bland Altman Limits of Agreement Plot. PwMS = people with multiple sclerosis, PwPD = people with Parkinson’s disease. Bias and upper and lower limits were calculated using all participants.

Figure 6 Stride Length Bland Altman Limits of Agreement Plot. PwMS = people with multiple sclerosis, PwPD = people with Parkinson’s disease. Bias and upper and lower limits were calculated using all participants.

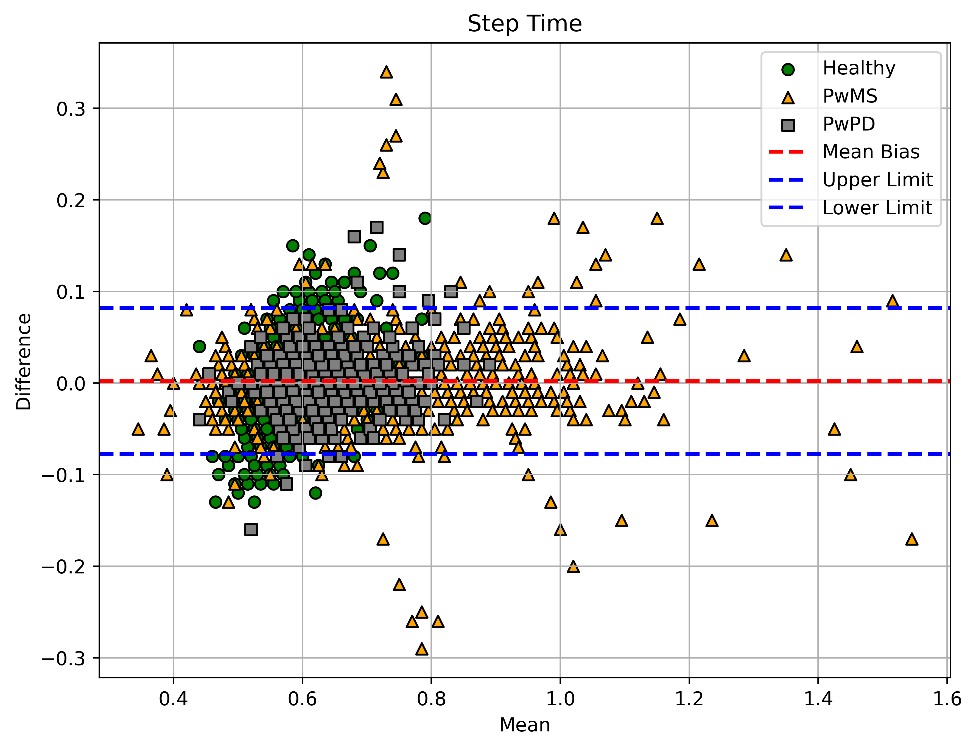

Figure 7 Step Time Bland Altman Limits of Agreement Plot. PwMS = people with multiple sclerosis, PwPD = people with Parkinson’s disease. Bias and upper and lower limits were calculated using all participants.

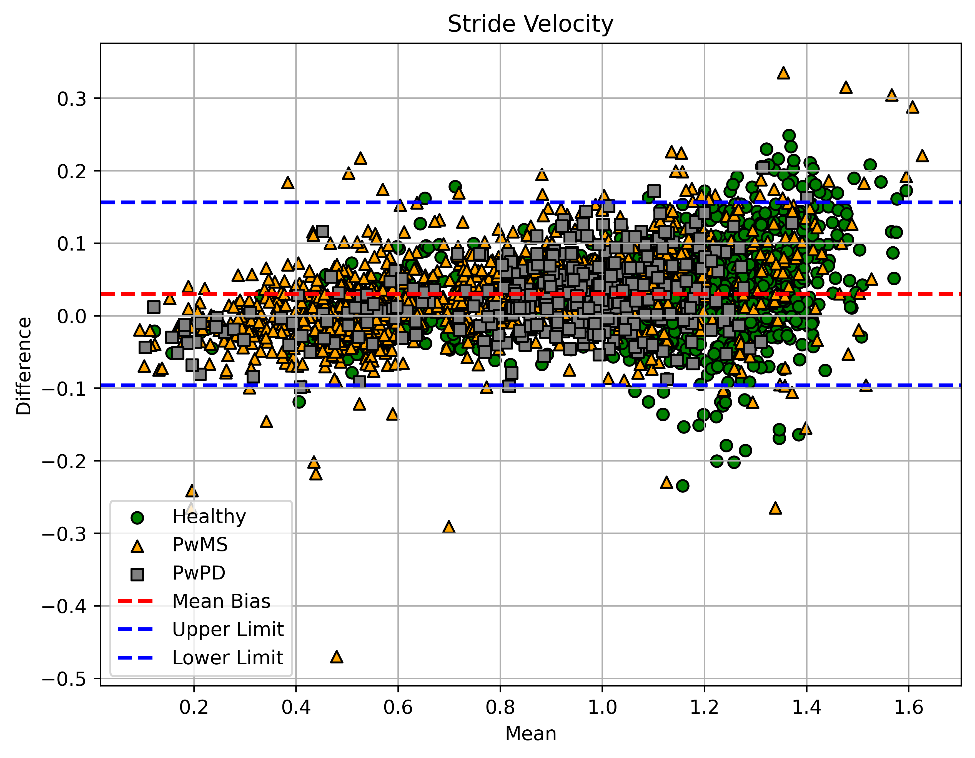

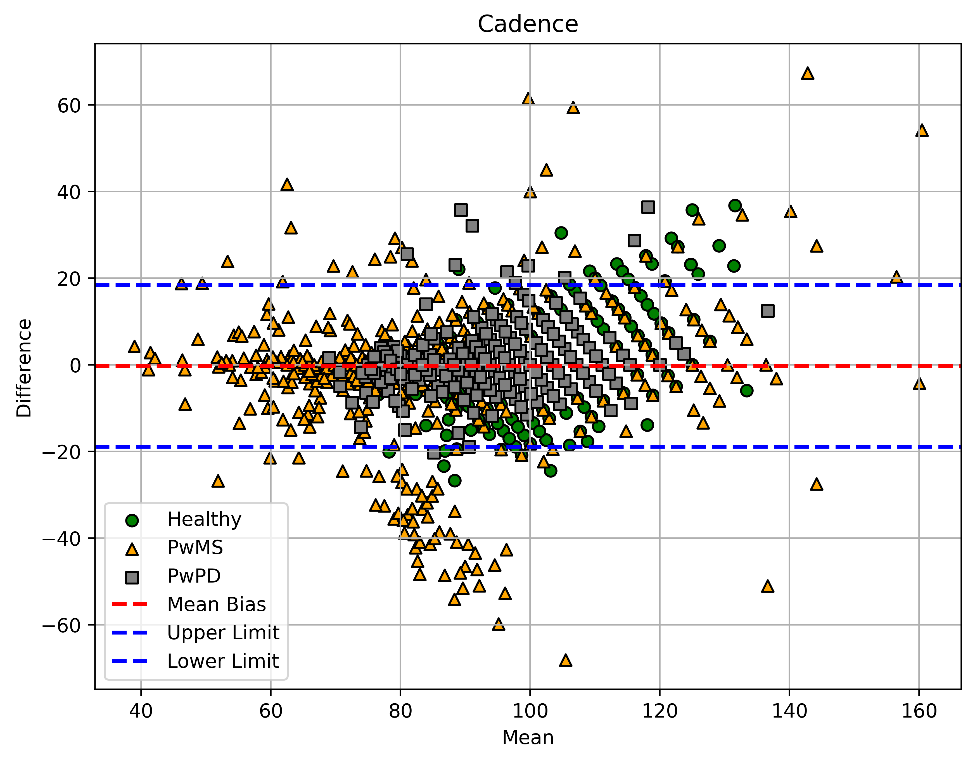

### Pace

Figure 8 Stride Velocity Bland Altman Limits of Agreement Plot. PwMS = people with multiple sclerosis, PwPD = people with Parkinson’s disease. Bias and upper and lower limits were calculated using all participants.

Figure 9 Cadence Bland Altman Limits of Agreement Plot. PwMS = people with multiple sclerosis, PwPD = people with Parkinson’s disease. Bias and upper and lower limits were calculated using all participants.

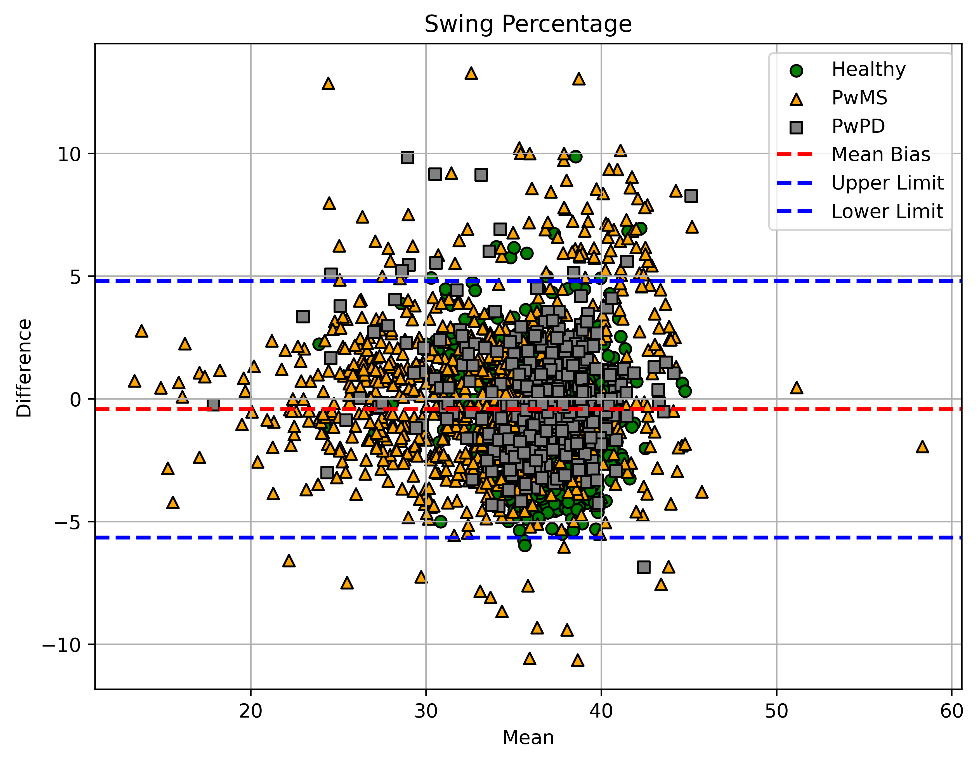

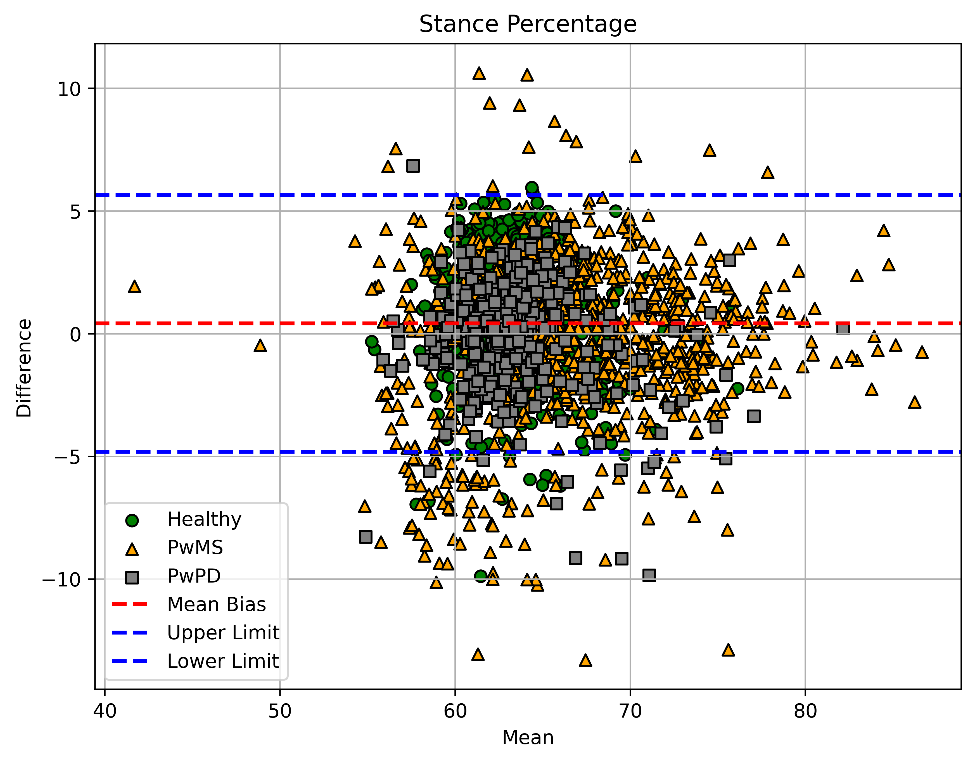

### Percentage

Figure 10 Stance Percentage Bland Altman Limits of Agreement Plot. PwMS = people with multiple sclerosis, PwPD = people with Parkinson’s disease. Bias and upper and lower limits were calculated using all participants. Values expressed as a percentage of Stride Time.

Figure 11 Swing Percentage Bland Altman Limits of Agreement Plot. PwMS = people with multiple sclerosis, PwPD = people with Parkinson’s disease. Bias and upper and lower limits were calculated using all participants. Values expressed as a percentage of Stride Time.

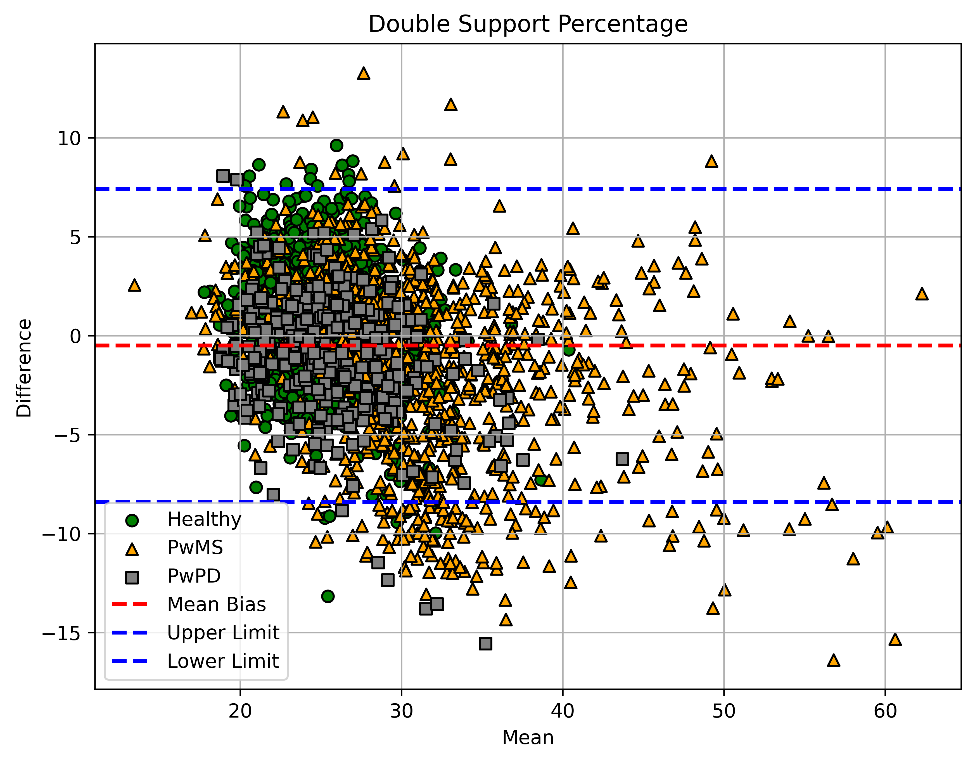

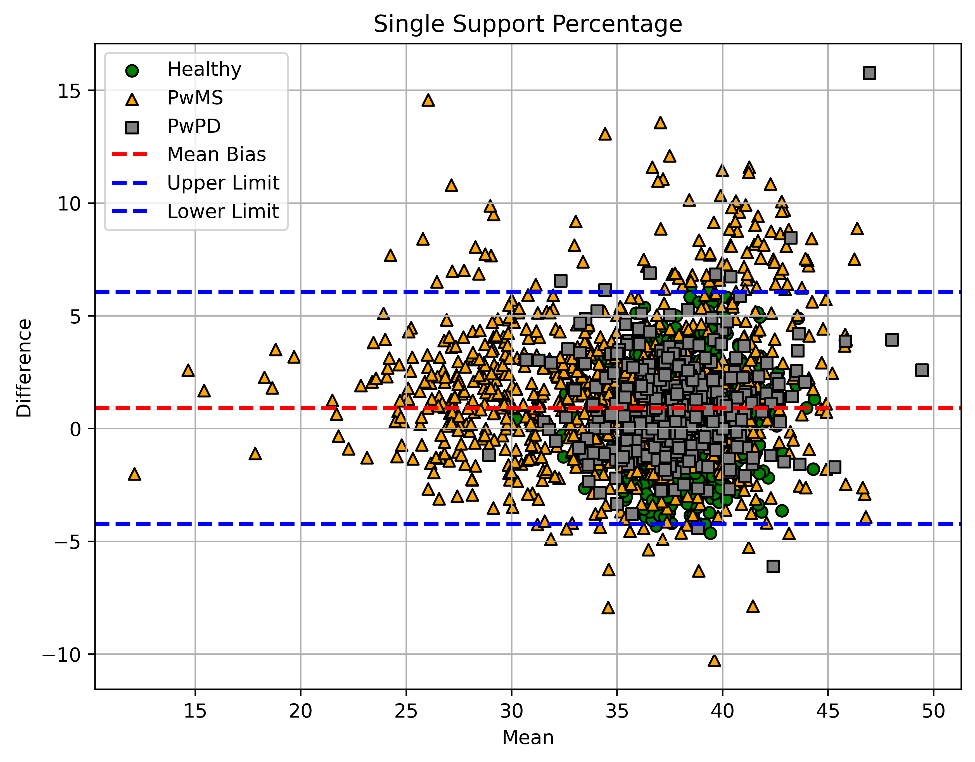

Figure 12 Single Support Percentage Bland Altman Limits of Agreement Plot. PwMS = people with multiple sclerosis, PwPD = people with Parkinson’s disease. Bias and upper and lower limits were calculated using all participants. Values expressed as a percentage of Stride Time.

Figure 13 Double Support Percentage Bland Altman Limits of Agreement Plot. PwMS = people with multiple sclerosis, PwPD = people with Parkinson’s disease. Bias and upper and lower limits were calculated using all participants. Values expressed as a percentage of Stride Time.

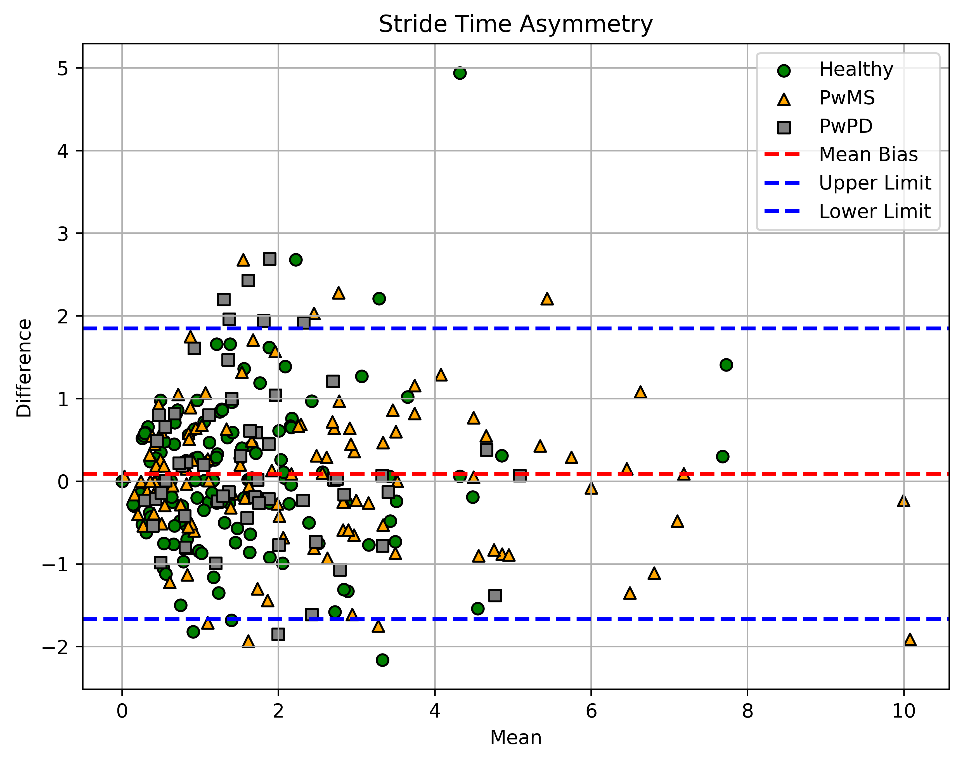

### Asymmetry

Figure 14 Stride Time Asymmetry Bland Altman Limits of Agreement Plot. PwMS = people with multiple sclerosis, PwPD = people with Parkinson’s disease. Bias and upper and lower limits were calculated using all participants. Values expressed as a percent difference between sides of the body.

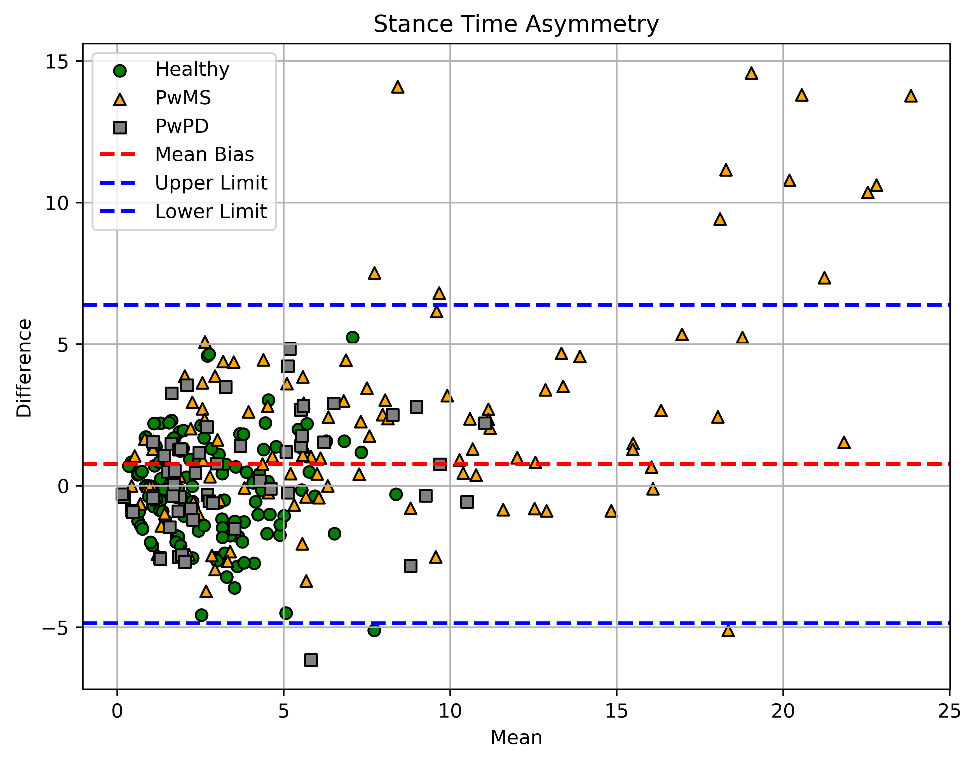

Figure 15 Stance Time Asymmetry Bland Altman Limits of Agreement Plot. PwMS = people with multiple sclerosis, PwPD = people with Parkinson’s disease. Bias and upper and lower limits were calculated using all participants. Values expressed as a percent difference between sides of the body.

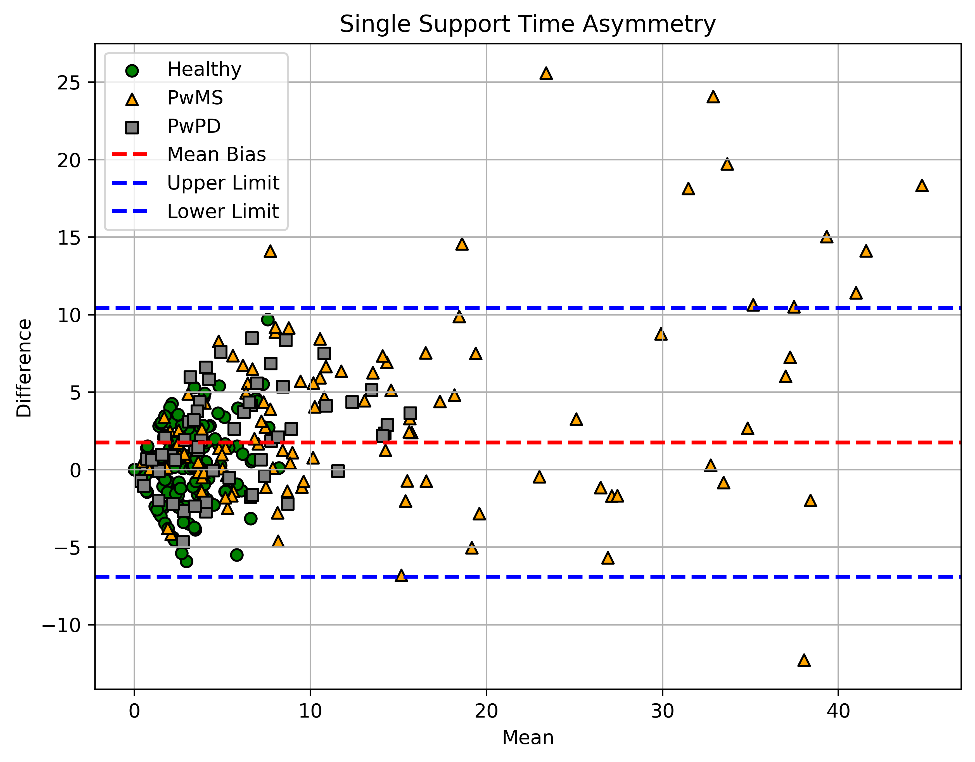

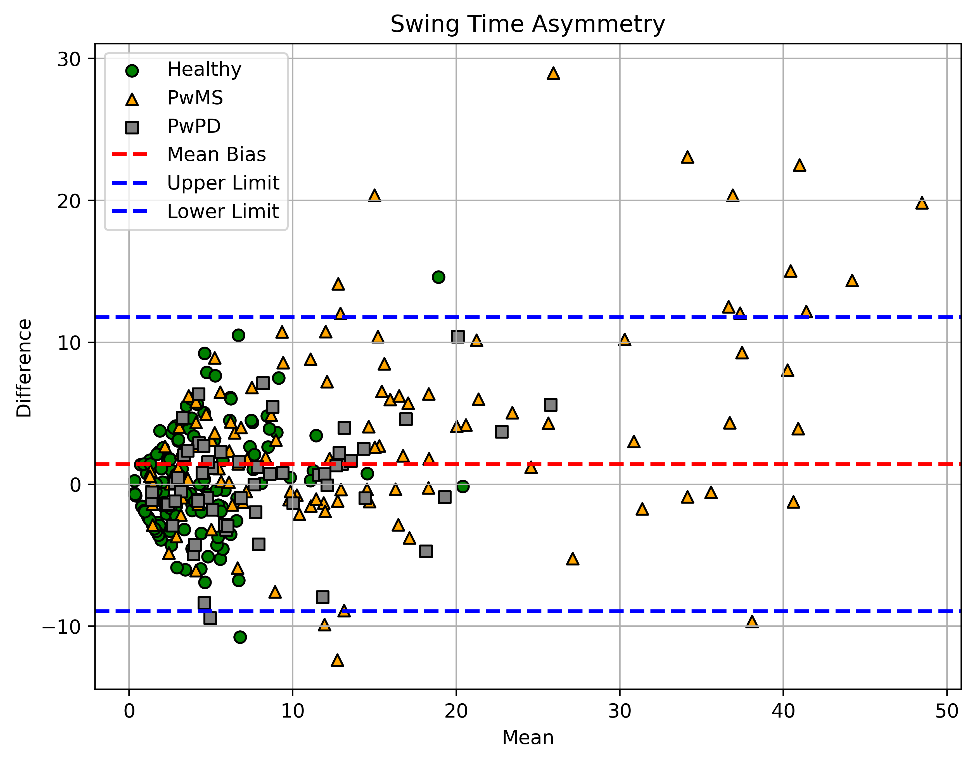

Figure 16 Swing Time Asymmetry Bland Altman Limits of Agreement Plot. PwMS = people with multiple sclerosis, PwPD = people with Parkinson’s disease. Bias and upper and lower limits were calculated using all participants. Values expressed as a percent difference between sides of the body.

Figure 17 Single Support Time Asymmetry Bland Altman Limits of Agreement Plot. PwMS = people with multiple sclerosis, PwPD = people with Parkinson’s disease. Bias and upper and lower limits were calculated using all participants. Values expressed as a percent difference between sides of the body.

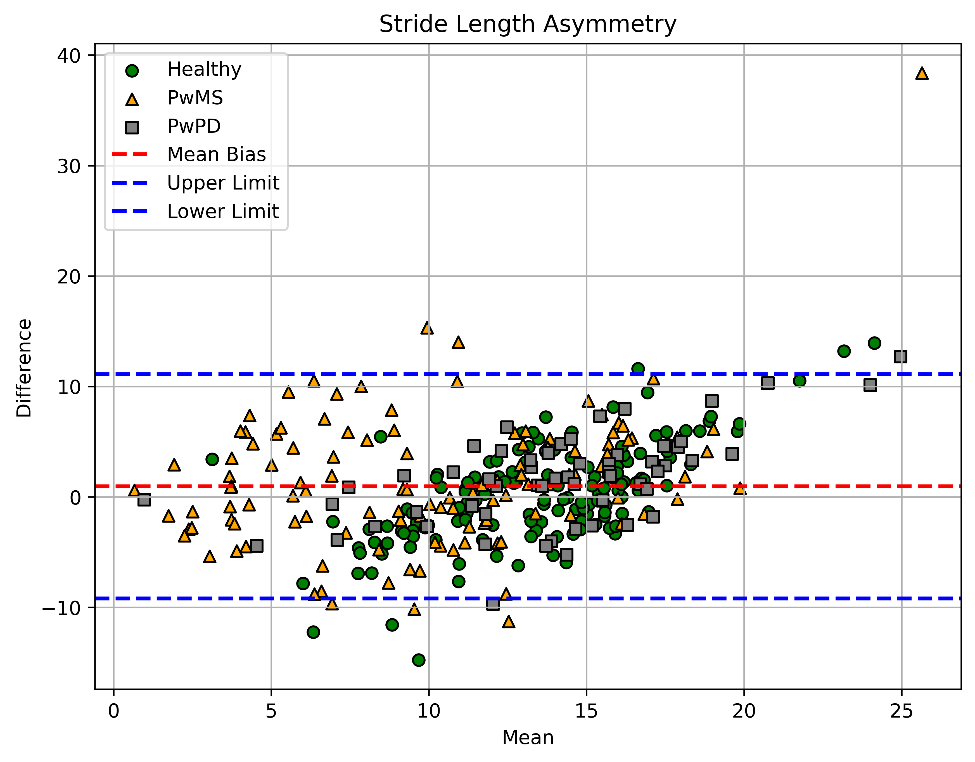

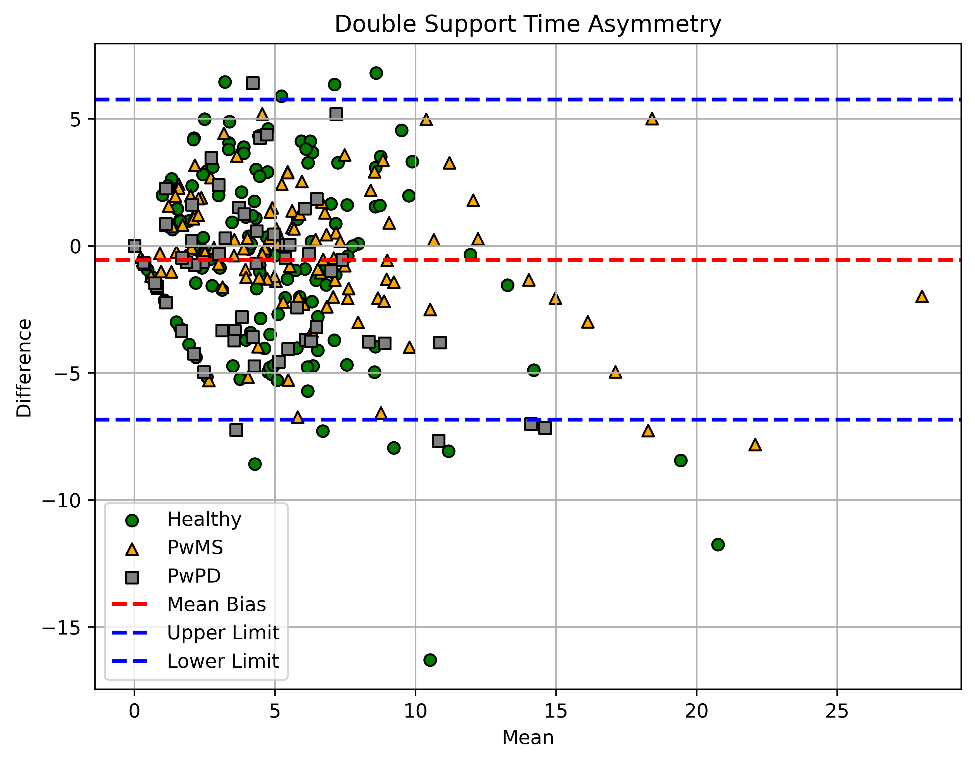

Figure 18 Double Support Time Asymmetry Bland Altman Limits of Agreement Plot. PwMS = people with multiple sclerosis, PwPD = people with Parkinson’s disease. Bias and upper and lower limits were calculated using all participants. Values expressed as a percent difference between sides of the body.

Figure 19 Stride Length Asymmetry Bland Altman Limits of Agreement Plot. PwMS = people with multiple sclerosis, PwPD = people with Parkinson’s disease. Bias and upper and lower limits were calculated using all participants. Values expressed as a percent difference between sides of the body.
