## Supplemental Variable Descriptions for "Validation of an instrumented shoe insole framework for analyzing spatiotemporal gait metrics in healthy and neurodegenerative populations"

Table S.1. Summary of spatiotemporal metric descriptions and calculations

| **Category** | **Metric** | **Description/Calculation** |
| --- | --- | --- |
| *Core* | Stride Time (s) | Time between heel strike events (same foot) |
|  | Stance Time (s) | Time between heel strike and toe off events (same foot) |
|  | Swing Time (s) | Time between toe off and heel strike events (same foot) |
|  | Single Support Time (s) | Time that only one foot is on the ground (same foot) |
|  | Double Support Time (s) | Time where two feet are on the ground |
|  | Stride Length (m) | Arclength distance between heel strike events (same foot) |
|  | Step Time (s) | Time between heel strike events of opposing feet |
| *Variability* | SD (individual core metrics; 7 metrics) | Standard deviation of each metric calculated in the ‘core’ category |
| *Pace* | Cadence (steps/min) | Step time × 60 s |
|  | Stride Velocity (m/s) | Stride length ÷ stride time |
| *Percentage* | Stance Percent (%) | $\left( \frac{Stance time}{Stride Time} \right)\times100$ |
|  | Swing Percent (%) | $\left( \frac{Swing time}{Stride Time} \right)\times100$ |
|  | Single Support Percent (%) | $\left( \frac{Single support time}{Stride Time} \right)\times100$ |
|  | Double Support Percent (%) | $\left( \frac{Double support time}{Stride Time} \right)\times100$ |
| *Asymmetry* | Stride Time Asymmetry (%) | $\left( \frac{\vert{Stride time}_{right}-{Stride time}_{left}\vert}{\left( {Stride time}_{right}-{Stride time}_{left} \right)\times0.5} \right)\times100$ |
|  | Stance Time Asymmetry (%) | $\left( \frac{\vert{Stance time}_{right}-{Stance time}_{left}\vert}{\left( {Stance time}_{right}-{Stance time}_{left} \right)\times0.5} \right)\times100$ |
|  | Swing Time Asymmetry (%) | $\left( \frac{\vert{Swing time}_{right}-{Swing time}_{left}\vert}{\left( {Swing time}_{right}-{Swing time}_{left} \right)\times0.5} \right)\times100$ |
|  | Stride Length Asymmetry (%) | $\left( \frac{\vert{Stride length}_{right}-{Stride length}_{left}\vert}{\left( {Stride length}_{right}-{Stride length}_{left} \right)\times0.5} \right)\times100$ |
|  | Single Support Time Asymmetry (%) | $\left( \frac{\vert{Single support time}_{right}-{Single support time}_{left}\vert}{\left( {Single support time}_{right}-{Single support time}_{left} \right)\times0.5} \right)\times100$ |
|  | Double Support Time Asymmetry (%) | $\left( \frac{\vert{Double support time}_{right}-{Double support time}_{left}\vert}{\left( {Double support time}_{right}-{Double support time}_{left} \right)\times0.5} \right)\times100$ |
